# Isolation of *Toxoplasma gondii*-specific IgG and mass spectrometric characterization of individual IgGs for early diagnosis of toxoplasmosis

**DOI:** 10.64898/2026.11.20.26367450

**Authors:** Mariama Souffou, Emmanuelle Demey, Magalie Dambrun, Alexandra Emmanuel, Zaineb Kammoun, Sébastien Dechavanne, Nawal Sare, Célia Dechavanne, Florence Migot-Nabias, Joëlle Vinh

## Abstract

**Objectives:** Toxoplasmosis represents a significant global health burden, particularly among immunocompromised patients and newborns of mothers who contracted a primary infection during pregnancy. In the newborn, maternally transferred *Toxoplasma gondii*-specific IgG persist for up to six months and mask the IgG newly synthesized by the infant, delaying diagnosis. Individual polymorphisms of the IgG heavy chain constant region (IGHG) can distinguish the two origins, but have so far been characterized only in total circulating IgG. We assessed whether this discrimination can be achieved on the IgG fraction directed against the parasite itself.

**Methods:** Recombinant *T. gondii* antigens (*Tg*SAG1, *Tg*GRA7, *Tg*AMA1) were produced and evaluated by ELISA on 85 seropositive and 95 seronegative plasma samples. Parasite-specific IgG were then immunopurified from a mixture of plasma from two donors, one seropositive and one seronegative, carrying distinct *IGHG1* and *IGHG3* alleles known from DNA sequencing, and their Fc/2 fragments were analyzed by middle-down and bottom-up LC-MS/MS.

**Results:** *Tg*SAG1 proved the most effective antigen; combined with *Tg*AMA1 and/or *Tg*GRA7 it reached 100% sensitivity and 97% specificity. Middle-down analysis offered high specificity but insufficient sensitivity for the microliter plasma volumes required in newborns. Bottom-up analysis identified the peptide WQQGNVFSCSVMHEGLHNHYTQK, characteristic of IGHG1*07, exclusively in the seropositive donor, in agreement with DNA sequencing data.

**Conclusion:** Combining antigen-affinity immunopurification with IGHG proteotyping allows an individual IGHG allele to be assigned within a pathogen-specific polyclonal IgG fraction representing less than 0.5% of total plasma IgG. This first demonstration, obtained in a two-donor context, opens the way to distinguishing maternal from fetal specific IgG for the early diagnosis of congenital toxoplasmosis.

## 1. INTRODUCTION

Toxoplasmosis is a cosmopolitan parasitosis that affects one-third of the world’s population [1]. This infection, caused by *Toxoplasma gondii*, is mostly asymptomatic, except in immunocompromised individuals and in fetuses and infants infected in utero. Congenital toxoplasmosis (CT) results from the transplacental passage of the tachyzoite blood stage of the parasite, which occurs in 30% of cases of primary infection during pregnancy [2]. The earlier the contamination of the fetus, the more serious the clinical consequences (death in utero, premature delivery, or term childbirth with perivisceral involvement), resulting mainly in neuro-ocular damage in case of later contamination [3,4]. Appropriate prenatal screening and treatment can significantly reduce the risks and severity of CT, which still presents a significant clinical challenge.

In countries that have chosen to implement prenatal toxoplasmosis monitoring, guidelines such as those established by the French National Authority for Health (HAS, Haute Autorité de Santé) [5] recommend monthly serological monitoring for *T. gondii* seronegative pregnant women to detect seroconversion by measuring immunoglobulin IgG and IgM levels [6]. However, in countries without automated screening and monitoring during pregnancy, the difficulty in diagnosing CT is exacerbated. This is the case on the African continent, where the toxoplasmosis prevalence rates fluctuate between 21%-74% depending on the region, with the highest rates in central African countries, and where no systematic recommendations are applied for pregnancy follow-up to prevent CT [7]. In France, the seroprevalence of toxoplasmosis in pregnant women has decreased over time, from 54.3% in 1995 to 31.3% in 2016, thanks to the national program established by the HAS for the prevention of congenital toxoplasmosis [6].

In addition to prenatal evaluation, neonatal biological diagnosis of toxoplasmosis is essential in the case of (i) suggestive clinical signs in the newborn with no information on the serological status of the mother, (ii) seroconversion diagnosed during pregnancy, (iii) not or poorly followed pregnancy, and (iv) for enhanced effectiveness of treatments administered as soon as possible to the newborn. This diagnosis is based mainly on the search for parasitic DNA in the placenta, amniotic fluid, cord blood, or newborn blood collected at delivery, in case of confirmed maternal seroconversion [6]. Serological tests are also performed on newborns. Indeed, as maternal IgA and IgM do not cross the placental barrier, they represent good markers of congenital infection in the newborn [8]. Nevertheless, some IgM could persist for years after the initial infection [9]. It has been demonstrated that IgM serology alone should no longer be considered an acute infection marker because of the heterogeneity of its kinetic response. [10] Detection of IgG synthesized by the child has diagnostic value only after 6 months of life, once maternally transmitted IgG has been eliminated. The immunoblotting techniques comparing the mother and child IgG profiles directed against several toxoplasmic antigens (Ags) remain difficult to interpret [8]. Nevertheless, the combination of several complementary tests constitutes a decision tree for a neonatal biological diagnosis of CT [6]. Early characterization of specific IgG neosynthesized by the newborn would be highly useful for diagnosing congenital *T. gondii* infection in the neonatal period.

To this end, a diagnostic test that differentiates maternal-specific IgG from newborn-synthesized IgG would be appropriate. The heavy chain (HC) of each IgG consists of a variable domain (VH) followed by three constant domains (CH1, CH2, and CH3-CHS), with a hinge region separating CH1 and CH2. The CH2 and CH3-CHS domains constitute the fragment crystallizable region or Fc region [11]. For each of the four IgG subclasses, IgG1, IgG2, IgG3, and IgG4, amino acid polymorphisms are described within the CH domains, mainly in the Fc fragments [12]. In the April 2025 edition, 73 alleles from the ImmunoGlobulin Heavy Gamma chain (*IGHG*) genes were referenced in the international database ImMunoGeneTics information system® (IMGT® [13], see S1 Table). They consist of 17 *IGHG1*, 19 *IGHG2*, 29 *IGHG3*, and 8 *IGHG4* alleles, respectively, for 6, 5, 18, and 5 distinct protein variants. To our knowledge, the exploration of their potential as a diagnostic tool in neonatal blood to distinguish pathogen-specific IgG transferred by the mother from that synthesized by the newborn in response to an in utero infection has not been documented. Our working group has progressively established the feasibility of this approach [14,15]. Dechavanne et al. first defined proteotypic peptides discriminating the G3m allotypes and the IGHG3 alleles, and showed that these could be detected and quantified by mass spectrometry in total IgG3 purified from plasma, including in a mother and her infant followed over the first months of life. Dambrun et al. subsequently extended the analysis to the four IGHG subclasses and validated it at the molecular level, alleles assigned by proteomics on total IgG matching those deduced from DNA sequencing of the CH2 and CH3-CHS exons in 95% of cases. A middle-down strategy applied to intact Fc/2 fragments of total polyclonal IgG further improved the specificity of allele discrimination [16]. All of this work was nevertheless carried out on the whole circulating IgG pool. Transposing it to a diagnostic setting requires isolating the IgG directed against the pathogen itself, a fraction amounting to less than 0.5% of total plasma IgG, and resolving the alleles of two individuals within a single sample. Unlike the characterization of monoclonal therapeutic antibodies (Abs), where a single IGHG variant is present at high concentration, the present work addresses a substantially more challenging analytical context. The simultaneous detection of up to eight co-occurring IGHG allelic variants in a polyclonal, Ag-specific IgG fraction representing less than 0.5% of total plasma IgG introduces near-isobaric signal overlap, severe sensitivity constraints, and allele-assignment ambiguities that require dedicated chromatographic and data-analysis strategies.

The isolation of plasma IgG specific to the *T. gondii* tachyzoite stage requires the use of the most adapted recombinant Ags for a diagnostic purpose in a CT context [17,18]. The *Tg*SAG1, *Tg*GRA7 and *Tg*AMA1 Ags have proven to be effective indicators of a *T. gondii* infection. Notably, dense granule Ags like GRA7 serve as specific markers for the acute phase of toxoplasmosis [19], while the major surface Ag SAG1 has shown high sensitivity in serology tests for both acute and chronic phases of the infection [20]. The *Tg*AMA1 apical membrane Ag is also investigated for its diagnostic potential, especially in the early infection stage [21].

In that context, the ultimate goal of our project is to provide an innovative approach to support an early diagnosis of congenital toxoplasmosis, based on the exclusive presence of *T. gondii-*specific IgG newly synthesized by the fetus, regardless of residual maternal IgG. This approach is based on a dual method that combines immunological isolation of mother- and child-*T. gondii*-specific plasma IgG, followed by mass spectrometry detection of the individual IGHG protein alleles. In the present study, we first evaluated the quality (specificity and sensitivity) of IgG directed against the selected *T. gondii* recombinant Ags (*Tg*SAG1, *Tg*GRA7 and *Tg*AMA1) using 180 plasma samples from a biobank of toxoplasmosis-seronegative and seropositive pregnant women. Second, we presented an intermediate proof-of-concept step of our alternative diagnostic approach, which consisted of using *T. gondii*-specific IgG isolated from 2 plasma donors, respectively seronegative and seropositive for toxoplasmosis, whose *IGHG* alleles had been determined using the DNA sequencing method described in [15]. A middle-down tandem MS protocol was designed exclusively for the proteomic assessment of these alleles, with a focus on the CH2 and CH3-CHS domains of the Fc fragment. We specifically further investigated the IgG1 and IgG3 subclasses by bottom-up proteomics for their prominent role in *T. gondii* congenital infection [22]. These two subclasses are all the more interesting because of the abundance of IgG1 (65 to 70% of total IgG) [23] on the one hand and the high amino acid polymorphism of IgG3 (18 protein variants) on the other hand.

## 2. MATERIALS AND METHODS

### 2.1 Biological samples

The study was performed using plasma samples obtained from pregnant women, seropositive or seronegative for *T. gondii*, to evaluate the specificity of the IgG response to the recombinant Ags produced. For this purpose, 180 plasmas, all preserved at -20°C until use and originating from three different field studies, were selected; their characteristics, including the recruitment periods, are presented in Table 1 [23, 24]. This enabled the collection of 95 seronegative and 85 seropositive plasma samples from different study designs and diagnosed by different methods for the present study. In addition, blood samples were collected from three healthy voluntary donors (HVD) on January 10, 2019. Their genomic DNA was used to characterize *IGHG* alleles following the DNA sequencing method described in [15]. Their plasma was divided into aliquots and stored at -20°C; these were then used for IgG processing, followed by mass spectrometry analysis. For the donor designated HVDtot, total IgG was enriched using Protein G. The other two donors were classified as either seropositive (HVD+) or seronegative (HVD-) for toxoplasmosis, based on the VIDAS® TOXO IgG II test (bioMérieux, France). Their IgG were immunopurified using affinity columns coated with *T. gondii* recombinant antigens. These two donors were selected because they carried distinct alleles of the *IGHG1* and *IGHG3* genes.

**Table 1.** Characteristics of the 3 population groups of pregnant women under study.

| **Origin of samples<sup>a</sup>** | **Inclusion dates** | **Location** | **Main objective** | **Serological diagnostic technique for toxoplasmosis** | **Toxoplasmosis serology** |  |
|---|---|---|---|---|---|---|
|  |  |  |  |  | **Pos. (n)** | **Neg. (n)** |
| **PlasDCty – PDC**<br>(*n = 57*, at delivery) | June 15, 2021 - November 15, 2021 | Akassato, Benin | Elucidation and circumvention of immune tolerance to malaria, with the aim of identifying a potential vaccine candidate that fully activates neonatal dendritic cells | Platelia™ Toxo IgG (Bio-Rad, France) | 27 | 30 |
| **CoaLa – COA**<br>(*n = 95*, at last prenatal visit) [24] | February 16, 2018 - April 9, 2018 | Cotonou, Benin | Comparison of the reliability of serological diagnostic tests for toxoplasmosis during pregnancy, based on the composition and combination of recombinant proteins used | recomWell IgG (Mikrogen Diagnostik, Germany) and confirmation with VIDAS® TOXO IgG II (bioMérieux, France) | 45 | 50 |
| **TOXODIAG – TXD**<br>(*n = 28*, at delivery) [25] | June 20, 2018 - December 12, 2022 | Ile de France, France | Characterization of the production of *Toxoplasma* antigen-specific immunoglobulins by neonatal B lymphocytes in the context of congenital infections | serological techniques used by the biology laboratories of the hospitals participating in the study | 13 | 15 |

<sup>a</sup>: The inclusion criteria for women were identical in all three studies: being at least 18 years old and giving birth without expected clinical complication.

### 2.2 Measurement of IgG specific to *T. gondii* recombinant antigens

*T. gondii* recombinant Ags, namely *Tg*AMA1, *Tg*SAG1 and *Tg*GRA7, were produced as described elsewhere [26] (see S1 File) and an ELISA was performed to measure the Ab response against them. Briefly, plates were coated with either purified human IgG (Sigma-Aldrich, St-Quentin-Fallavier, France) at concentrations ranging from 10 to 5000 ng/ml, to perform a standard range, or with each recombinant Ag at 1.5 µg/ml in 1X PBS coating buffer. Plasma samples were diluted to 1:100 in dilution buffer. Each sample was tested in duplicate. The concentration of specific Abs was calculated using the free online analysis software Myassays (https://www.myassays.com/). For each IgG response, the cut-point was calculated using a Receiver Operating Characteristic (ROC) curve with a confidence interval for the area under the ROC curve (AUC), and a p-value ≤ 0.05 was considered statistically significant. The ROC curves were generated for each Ag analyzed using GraphPad Prism software (v.10).

### 2.3 Isolation of *T. gondii-*specific IgG on columns coated with monoclonal anti-His magnetic beads

This part was carried out using plasma from 2 voluntary donors, who were seronegative (HVD-) and seropositive (HVD+) for toxoplasmosis, respectively, and whose IGHG protein alleles had been deduced using the DNA sequencing method described in [15]. Ag-specific IgG was isolated separately for each of the two-donor sample to assess the specificity of the affinity column. Subsequently, equal volumes of plasma from *T. gondii* negative and positive donors were mixed and Ag-specific IgG was purified from the resulting plasma mixture. For this purpose, magnetic beads covalently coated with monoclonal anti-His were used (anti-His Magnetic Beads, CliniSciences, France). The binding capacity of the beads was over 0.5 mg/ml of 6 His-tagged proteins. An adjusted volume of beads was added to the column to reach an equimolar Ag concentration of 48.34 nM in a total volume of 10 ml of PBS 1X at pH 9 (S2 Table). *Tg*AMA1, *Tg*SAG1 and *Tg*GRA7 recombinant proteins were coated separately in columns and stirred overnight at +4°C. The Ag-coated columns were then placed on a magnetic support to remove the supernatant. The coated beads were washed and blocked with PBS 1X - 0.5%Tween 20 - 3% BSA for 1h at +4°C. Plasma samples were diluted at 1:150 in 10 mL of dilution buffer (PBS 1X - 0.5% Tween 20 - 1% BSA - 0.02% sodium azide) and incubated with each *T. gondii* Ag in each column for 1h30 to allow specific Ag-Ab interaction. The columns were then washed twice with PBS 1X - 0.5% Tween 20 - 0.5 M NaCl, once with PBS 1X - 0.5% Tween 20 - 0.5 M NaCl - 0,5 M urea, then once with PBS without detergent. Finally, specific IgGs were eluted in PBS-1% formic acid and immediately neutralized with 1 M Tris at pH 9 to prevent protein denaturation. The quality of isolated *T. gondii-*specific IgG was verified by Western blot utilizing an HRP-conjugated anti-human Fc antibody (Jackson ImmunoResearch Europe, St. Thomas’ Place, UK) diluted at 1:5000.

### 2.4 Isolation of Fc/2 fragments from *T. gondii*-specific IgG subclasses using nanobodies

The three eluted fractions obtained after purification Ag-specific IgG from the mixture of *T. gondii* negative and positive plasma samples were pooled then fractionated by successive passages over Capture Select™ resin specific for each IgG to allow enrichment in Fc: after a first incubation with 20 µl of Capture Select™ IgG3 (Hu) Affinity Matrix (ThermoFisher Scientific) (previously washed with 1X PBS, pH 7) for 60min in 1X PBS at room temperature (binding phase), the mixture was centrifuged (12,000xg, 5min). The supernatant from the Capture Select™ IgG3 incubation was collected and subsequently incubated with Capture Select™ IgG1 (Hu) Affinity Matrix (ThermoFisher Scientific) using the same protocol. The affinity resins were washed (twice with 500 µl PBS 1X, and twice with 500 µl of purified water), and the specific IgG subclasses were eluted in both fractions by adding 40 µl of 1% formic acid (low pH elution buffer), followed by neutralization with 40 µl of 1.5 M Tris-HCl pH 8.8. Protein content was assessed using a 2 µl aliquot using a NanoDrop (ThermoFisher Scientific, Waltham, MA, USA).

#### 2.4.1 PNGase F deglycosylation

1U of active PNGase F (Promega) per 10 µg of IgG (as estimated by NanoDrop) was added to 80 µl of the Capture IgG1 and Capture IgG3 eluate solutions. Each mixture was incubated at 37°C for 18 hours and then cooled to room temperature before proceeding to the next step.

#### 2.4.2 IdeS digestion

IdeS (Genovis), proteinase from *Streptococcus pyogenes*, cleaves IgG antibodies at the lower hinge region with high specificity between two glycine residues. This cleavage results in the generation of an F(ab′)2 fragment and an Fc fragment. To maintain the specificity of capture by the Capture Select affinity resin, the reaction must be performed after isolation of IgG1 and IgG3. A diluted IdeS enzyme solution at 6.7 U/µl was added to the two deglycosylated fractions, with 1 U of IdeS acting on 1 µg of IgG, followed by incubation at 37°C for 1 hour.

#### 2.4.3 Disulfide bond reduction

A sufficient amount of 50 mM TCEP (Tris(2-carboxyethyl)phosphine hydrochloride, ThermoFisher Scientific) diluted in 1X PBS to achieve a final concentration of 10 mM TCEP in 1X PBS was added to each fraction, followed by incubation at 37°C for 2 hours. The reduction generates three Ab fragments: two Fd heavy chain fragments, two Lc light-chain fragments, and two Fc/2 fragments containing the targeted CH2 and CH3-CHS Fc domains.

#### 2.4.4 Fc/2 capture

Finally, the different fractions were incubated on Capture Select FcXP (Affinity Matrix, ThermoFisher Scientific), washed, eluted, and neutralized using the protocol described above to isolate the Fc domains. This yielded two negative control fractions, FcX3(-) and FcX1(-), and two *Toxoplasma*-positive fractions, FcX3(+) and FcX1(+).

### 2.5 Mass spectrometry characterization of IGHG alleles from Fc/2 amino-acid sequences

We briefly described the different steps. A detailed protocol is provided in S1 File.

#### 2.5.1 Middle-down analysis

IGHG alleles were detected on a Q Exactive HF Orbitrap coupled with a nanoRSLC system. Samples were separated on a C4 column and acquired in DDA mode (240,000 resolution; top 10 HCD fragmentation). Spectra were deconvoluted using Freestyle 1.8 and searched with Byonic against a custom IMGT-derived IGHG library (10 ppm precursor tolerance; filters: score >250, False Discovery Rate <1%). Results were manually validated.

#### 2.5.2 IGHG sequence library

A reference database was built from IMGT FASTA sequences covering CH2–CH3-CHS regions of IGHG1–4 completed with the amino-acid sequences of Khatri et al. [27]. Full CH1-to-M2 coding sequences were included to ensure complete coverage of all proteolytic peptides. One previously identified unreferenced IGHG1 allele was also added.

#### 2.5.3 Bottom-up sample preparation

Fc/2-containing fractions were reduced with 5 mM DTT for 30 min at 56°C, alkylated with 25 mM iodoacetamide at room temperature in the dark, and digested overnight at 37°C with trypsin, LysC, or both. Peptides were desalted with ZipTip C18 (Millipore, Merck).

#### 2.5.4 Fc/2 chromatographic separation

After evaluating C4, MabPac RP, and Poroshell 300SB-C3 stationary phases, the latter was selected and connected on a UHPLC Vanquish system (80 μL/min, 60°C) coupled to an Orbitrap Tribrid Eclipse as the operational middle-down platform.

#### 2.5.5 DDA and DIA bottom-up proteomics

Peptides were injected onto a Neo Vanquish nanoLC coupled to an Orbitrap Astral. Separation was performed on a 25 cm C18 Aurora column using a 40-min gradient. DDA was run in Top100 mode; DIA used 2 m/z windows over 450–1500 m/z. Identification was performed using Sequest-HT (DDA) and DIA-NN (DIA), with carbamidomethylation fixed and oxidation/deamidation variable. Candidate allele-specific peptides were verified at the MS1 and MS2 levels with 30 ppm tolerance.

### 2.6 Statistical analysis

The performance of the three recombinant Ags in terms of sensitivity (SS) and specificity (SP) of IgG responses directed against them was determined from serological results for the three investigated population groups.

#### 2.6.1 ROC curves and cut-point optimization

The ROC curve analysis evaluates the SS and SP variations of each Ag tested by ELISA. A curve closer to the upper left corner of the graph indicated better discrimination between toxoplasmosis seropositive and seronegative groups [28]. The AUC value reflected the test’s diagnostic accuracy. The Youden Index (*J*) identified the optimal cut-point, corresponding to the maximum index value achieved, optimizing the discriminatory power of the Ags [29]. According to the ROC curve analyses, quantitative results obtained from ELISA were dichotomized into positive (1) and negative (0) IgG responders to each recombinant Ag.

#### 2.6.2 Validation of the diagnostic performance of the *T. gondii* recombinant antigens

SS and SP were calculated after applying the optimal cut-point to seropositive (n = 85) and seronegative (n = 95) samples, respectively. The SS represents the percentage of true positives (TP) correctly identified in response to each recombinant Ag among all toxoplasmosis seropositive samples, including false negatives (FN). SP indicates the proportion of true negatives (TN) individuals correctly detected in the toxoplasmosis seronegative samples (n=95), including false positives (FP).

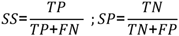

In addition, the Positive Predicted Value (PPV) and the Negative Predicted Value (NPV) indicate the likelihood that the probability that a positive or negative result is accurate [30].

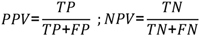

### 2.7 Ethics

The TOXODIAG study “New diagnostic approach for congenital toxoplasmosis” received the ethical clearance of the Comité de Protection des Personnes (CPP) Sud-Méditerranée II from France on October 3, 2017, under the ID RCB 2017-A02208-45. The CoaLa study “Caractérisation de la production d’immunOglobulines spécifiques d’antigènes pArasitaires par les Lymphocytes B néonAtaux dans le cadre d’infections congénitales” was authorized by the Comité National d’Ethique pour la Recherche en Santé (CNERS) from Benin, with the n°40 assigned on September 28, 2017. Similarly, the PlasDCty study “Elucider et contourner la tolérance immunitaire au paludisme” was authorized by the CNERS under the n°4 allocated on March 23, 2021. For each study, enrolled women provided written informed consent before their inclusion and approved the use of the collected data. This also applied to the three voluntary donors, who were recruited on January 10, 2019. All individuals were explicitly given the choice to withdraw from each study at any stage. Each study adhered to the WMA Declaration of Helsinki, adhering to ethical principles for medical research involving human subjects. All the methods were performed in accordance with the approved guidelines and regulations.

## 3. RESULTS

### 3.1 *Tg*SAG1 recombinant antigen shows higher diagnostic accuracy than *Tg*AMA1 and *Tg*GRA7

IgG responses against each recombinant Ag were measured in all individuals, grouped by toxoplasmosis serological status (Fig 1). ROC curve analysis assessed the diagnostic performance of each Ag for distinguishing seropositive from seronegative samples (Fig 2) [28]. This revealed that *Tg*SAG1 had the best diagnostic performance with an AUC of 0.99 (p<0.0001), indicating near-perfect sensitivity and specificity. This suggests that *Tg*SAG1 is highly accurate at discriminating between positive and negative samples. With an AUC of 0.93 (p< 0.0001), *Tg*AMA1 also demonstrated a highly effective diagnostic value, greater than that of *Tg*GRA7 (AUC of 0.85, p< 0.0001), indicating less sensitivity and specificity but still acceptable diagnostic performance.

**Fig 1.**
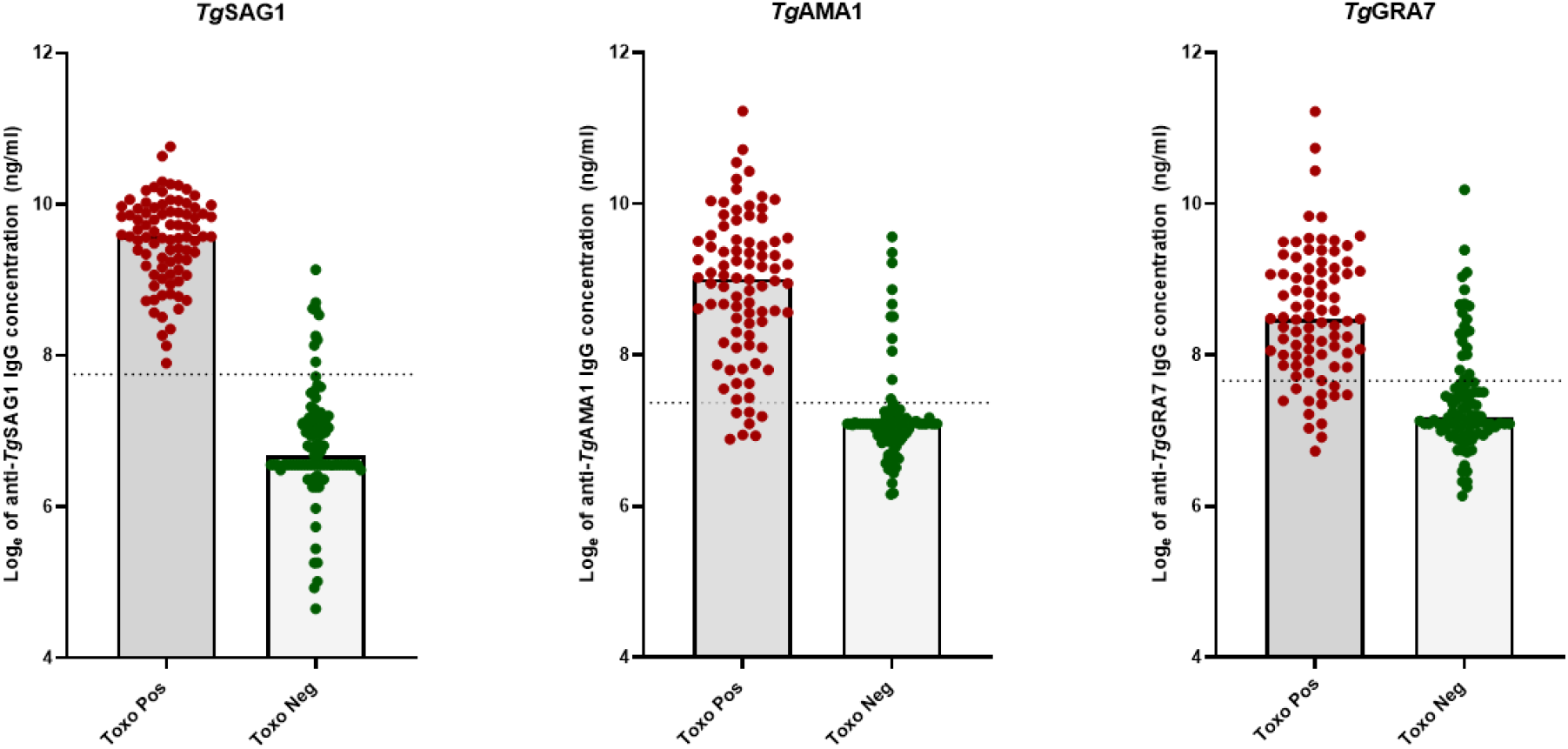
Distribution of the individual IgG responses against *Tg*SAG1, *Tg*AMA1 and *Tg*GRA7 in the toxoplasmosis seropositive (n=85) and seronegative (n=95) groups. The histogram bar in the scatter dot plot represents the median and the dot line represents the optimal cut-point. Green dots represent seronegative individuals. Red dots represent seropositive individuals. For the graphical design, we converted IgG concentrations measured in ng/mL to logarithm base e (natural logarithm). The dashed line represents the optimal cut-point determined by the Youden index (see Table 2).

**Fig 2.**
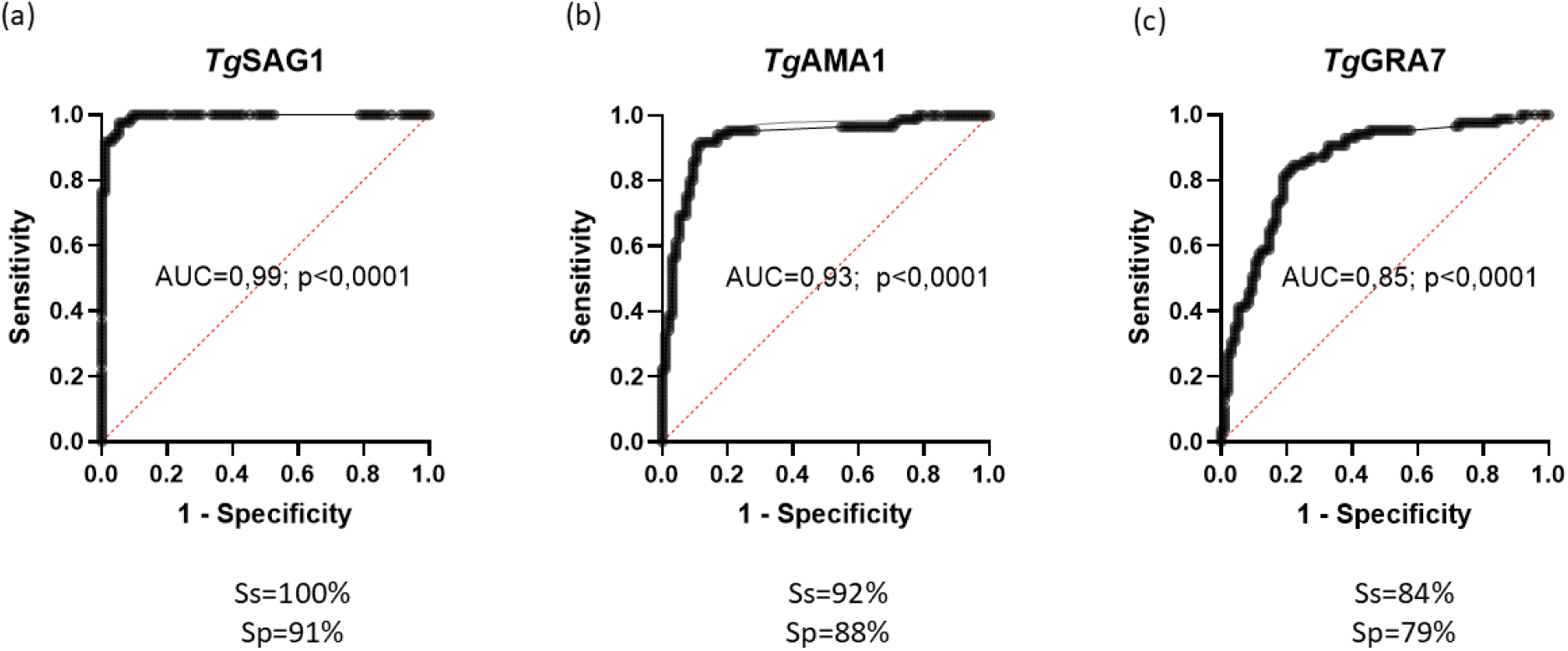
ROC curves for the evaluation of the diagnostic performances of *Tg*SAG1, *Tg*AMA1 and *Tg*GRA7 recombinant antigens. **(a)** *Tg*SAG1 achieved 100% sensitivity and 91% specificity at the optimal cut-point; **(b)** *Tg*AMA1 achieved 92% sensitivity and 88% specificity at the optimal cut-point; **(c)** *Tg*GRA7 achieved 84% sensitivity and 79% specificity at the optimal cut-point. The diagonal reference line of the ROC curve represents a region of complete randomness of the test (AUC = 0.5), which does not differentiate between *T. gondii* seropositive (n=85) and seronegative (n=95) samples. The AUC ranges from 0 to 1, with a value of 0 indicating a totally inaccurate test and a value of 1 indicating a perfectly accurate test. Generally, an AUC of 0.5 suggests no discrimination, 0.7 to 0.8 is considered acceptable, 0.8 to 0.9 is considered excellent, and >0.9 is considered outstanding [28].

**Table 2.**
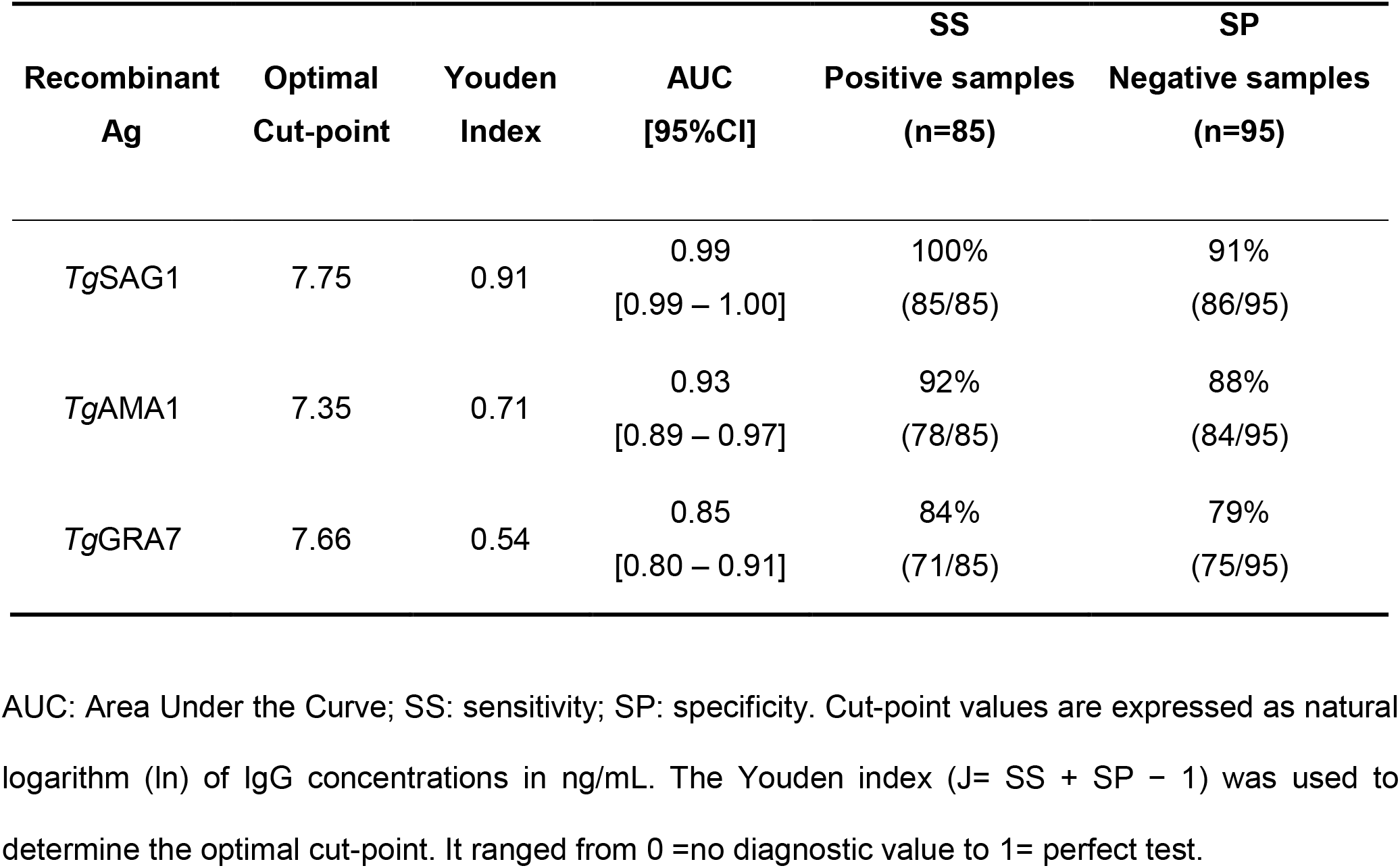
Performance of each *T. gondii* recombinant antigen according to the ROC characteristics and the Youden index.

In addition to AUC values, the Youden Index is a comprehensive measure of the test’s effectiveness, combining sensitivity and specificity to help determine the optimal cut-point that maximizes a test’s performance. For *Tg*SAG1, the optimal cut-point was 7.75, with a high Youden Index of 0.91, indicating an excellent overall performance (Table 2). The test achieved 91% specificity, identifying the majority of true negatives, and 100% sensitivity, detecting all true positives accurately. Indeed, Fig 1 shows that seronegative individuals (green dots) had Ab concentrations mainly below the threshold, whereas all seropositive individuals (red dots) had Ab concentrations well beyond the detection threshold. For *Tg*AMA1, the best cut-point was 7.35, and its moderate Youden Index of 0.71 indicated a respectable test performance (Table 2). With 92% sensitivity and 88% specificity, *Tg*AMA1 showed very good performance; however, it was marginally lower than that of *Tg*SAG1. This pattern reflected the individual results shown in Fig 1, in which the IgG responses of most seronegative or seropositive women were below or above the threshold, respectively. Lastly, *Tg*GRA7 exhibited a moderate performance with an optimal cut-point of 7.66 and a reduced Youden Index of 0.54 (Table 2). It was the least useful of the three Ags for diagnostic purposes, performing as well as *Tg*SAG1 and *Tg*AMA1 but with less consistency (84% sensitivity and 79% specificity), as shown with the individual distribution of values (Fig 1).

### 3.2 The combination of the individual antibody response against *Tg*SAG1 with *Tg*AMA1 and/or *Tg*GRA7 is the most performant for the serological diagnosis of toxoplasmosis

The diagnostic performance of different combinations of IgG responses against recombinant Ags was analyzed in order to determine the optimal Ag combination regarding the sensitivity, specificity, positive and negative predictive values of the Ab responses.

The [*Tg*SAG1+*Tg*AMA1] combined IgG responses exhibited a sensitivity of 16%, meaning that only 14 among the 85 toxoplasmosis-positive samples responded positively to *Tg*SAG1 and *Tg*AMA1 (Table 3). This combination also had a perfect PPV of 100%, indicating complete reliability in positive results, but the PNV was moderate at 57%, suggesting limited ability to identify negatives correctly. The combination of positive IgG responses against [*Tg*SAG1+*Tg*AMA1+*Tg*GRA7] achieved a much higher sensitivity of 75%, thereby improving the detection of true positives. It also had an excellent specificity of 99%. Combining the 3 Ags yielded a PPV of 98% and a PNV of 82%, demonstrating that it performed better than [*Tg*SAG1+*Tg*AMA1] or [*Tg*SAG1+*Tg*GRA7] only (Table 3). However, the [*Tg*SAG1+*Tg*XrecAg] combination, which is the association of a positive IgG response to *Tg*SAG1 with another one, directed to *Tg*AMA1 or *Tg*GRA7 or both, outperformed all the others, showing not only a 100% sensitivity and a PPV of 97%, but also a specificity of 97% with a PNV at 100%, thus making this combination the most reliable in both recognizing true positives and true negatives. Interestingly, the combination [*Tg*AMA1+*Tg*GRA7] and the isolated responses to only one Ag, either *Tg*SAG1, *Tg*AMA1 or *Tg*GRA7, were unreliable, with sensitivity and PPV both at 0%, making them worthless for recognizing true positives, showing the importance of combining Ags with at least *Tg*SAG1 (Table 3).

**Table 3.** Evaluation of the combined IgG responses against several *T. gondii* recombinant antigens.

| **Combined IgG antibody responses to several *Tg* Ags** | **SS<br>Positive samples<br>(n=85)** | **SP<br>Negative samples<br>(n=95)** | **PPV** | **NPV** |
|---|---|---|---|---|
| [*Tg*SAG1+*Tg*AMA1] | 16%<br>(14/85) | 100%<br>(95/95) | 100%<br>(14/14) | 57%<br>(95/166) |
| [*Tg*SAG1+*Tg*GRA7] | 8%<br>(7/85) | 98%<br>(93/95) | 78%<br>(7/9) | 54%<br>(93/171) |
| [*Tg*SAG1+*Tg*AMA1+*Tg*GRA7] | 75%<br>(64/85) | 99%<br>(94/95) | 98%<br>(64/65) | 82%<br>(94/115) |
| **[*Tg*SAG1 + *Tg(*X)recAg]** | **100%**<br>(85/85) | **97%**<br>(92/95) | **97%**<br>(85/88) | **100%**<br>(92/92) |
| [*Tg*AMA1+*Tg*GRA7] | 0%<br>(0/85) | 95%<br>(90/95) | 0%<br>(0/5) | 51%<br>(90/175) |
| [*Tg*SAG1/*Tg*AMA1/*Tg*GRA7] | 0%<br>(0/85) | 76%<br>(72/95) | 0%<br>(0/23) | 46%<br>(72/157) |

[*Tg*SAG1+*Tg*AMA1]: combined IgG responses to *Tg*SAG1 and *Tg*AMA1. [*Tg*SAG1+*Tg*GRA7]: combined IgG responses to *Tg*SAG1 and *Tg*GRA7. [*Tg*SAG1+*Tg*AMA1+*Tg*GRA7]: combined IgG responses to all three recombinant Ags.
**[*Tg*SAG1 + *Tg*(X)recAg]**: IgG response against *Tg*SAG1 combined with that directed to either *Tg*AMA1 or *Tg*GRA7 or both *Tg*AMA1 and *Tg*GRA7. [*Tg*AMA1+*Tg*GRA7]: combined IgG responses to *Tg*AMA1 and *Tg*GRA7.
[*Tg*SAG1/*Tg*AMA1/*Tg*GRA7]: IgG response to only one recombinant Ag. Among the seropositive samples, none of the 85 individuals responded to a single Ag, all showed responses to at least two Ags, always including *Tg*SAG1.
Among the 95 negative samples, the 23 false-positive individuals responded to only one Ag (*Tg*SAG1 = 6, *Tg*AMA1 = 5 and *Tg*GRA7 = 12). PPV (Positive Predictive Value) et NPV (Negative Predictive Value).

### 3.3 *T. gondii*-specific IgG isolated from a seropositive sample bind specifically to *T. gondii* recombinant antigens

This experiment was carried out with the 2 samples, HVD- and HVD+. The IgG directed to *Tg*SAG1, *Tg*AMA1, and *Tg*GRA7 recombinant Ags were isolated using columns coated with monoclonal anti-His magnetic beads. Fig 3a shows that these IgG bind to each Ag only in the seropositive sample. Indeed, distinct bands matching the human heavy chain weight (50 kDa) were observed, indicating the presence of specific IgG, with a strong response to *Tg*SAG1. Conversely, the seronegative sample shows few discernible bands, suggesting a background of residual IgG. These results were corroborated by the immunoblot profile of the IgG eluted fractions obtained after mixing equal volumes of plasma from the *T. gondii*-seropositive individual (HVD+) and the seronegative individual (HVD-) prior to the purification of Ag-specific IgG. The immunoblot profile obtained from the IgG purification of the mixed plasma was similar to that of the purified IgG from the seropositive sample alone. This finding indicates that the presence of IgG from the seronegative plasma does not interfere with the specific Ag–Ab interaction (Fig 3b). These findings support the specificity of the recombinant Ags in identifying *T. gondii*-specific Abs in seropositive samples.

**Fig 3.**
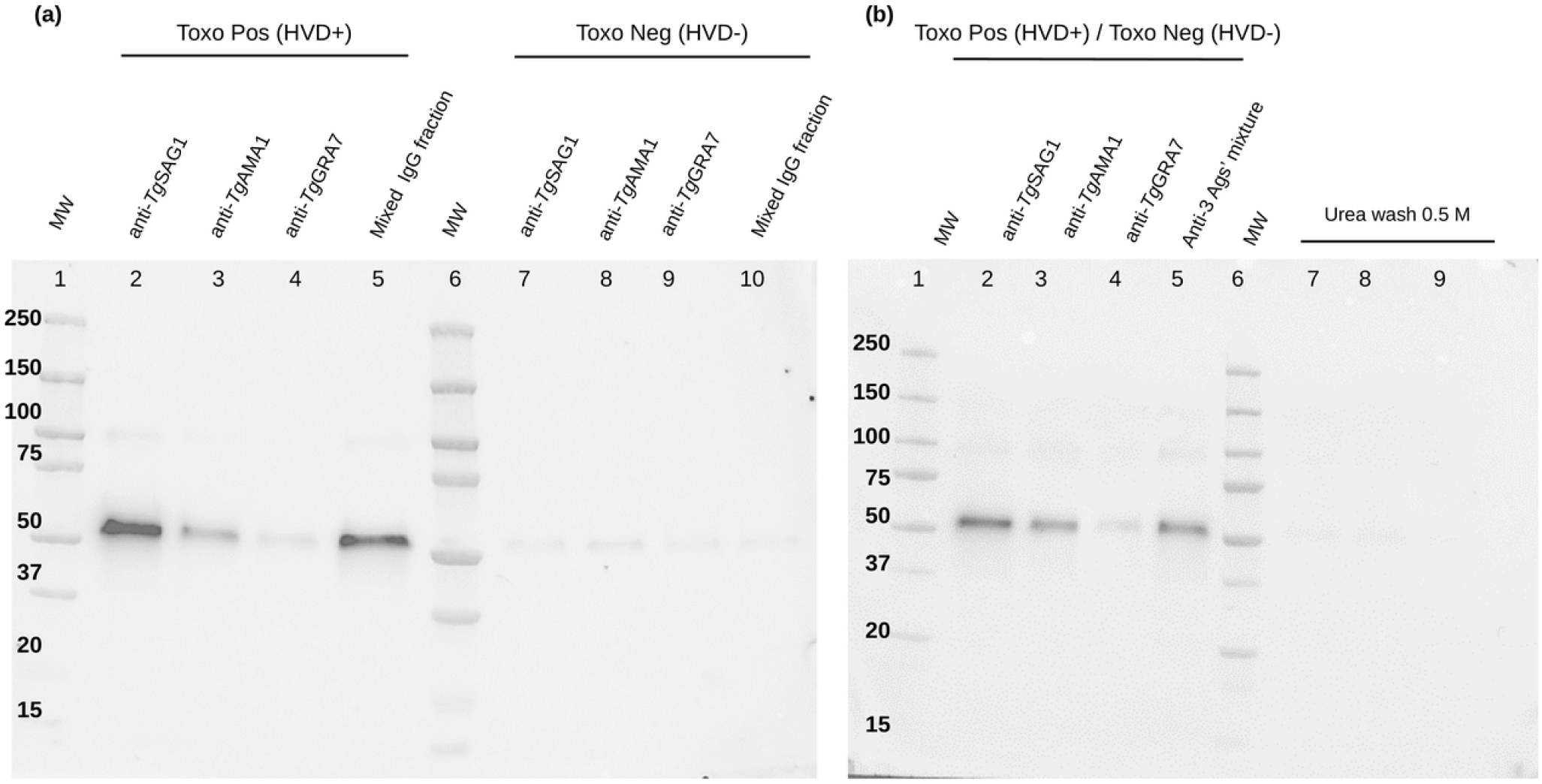
Recognition of the produced *Tg*SAG1, *Tg*AMA1 and *Tg*GRA7 recombinant antigens by IgG purified from *T. gondii* seronegative and seropositive samples (Western blot). **(a)** Toxo Pos (HVD+): plasma sample seropositive for toxoplasmosis; Toxo Neg (HVD-): plasma sample toxoplasmosis seronegative. Lanes 2 to 4 correspond to the specific IgG fractions directed against the recombinant antigens *Tg*SAG1, *Tg*AMA1 and *Tg*GRA7, respectively, from HVD+. Lanes 7 to 9 correspond to the antigen-specific IgG fractions from HVD-. Lane 5: the mixed IgG fraction corresponds to a mixture of the IgG fractions directed against each antigen, in equal volume for the seropositive sample (lanes 2 to 4) and the seronegative sample (fractions 7 to 9). **(b)** Toxo Pos (HVD+) / Toxo Neg (HVD-): an equal-volume mixture of plasma samples seropositive and seronegative for toxoplasmosis, respectively. Lanes 2 to 4 show the cumulative IgG responses against the recombinant antigens *Tg*SAG1, *Tg*AMA1, and *Tg*GRA7, respectively. Lane 5 shows the cumulative IgG response against a mixture of the three recombinant antigens combined in equal volumes. Lanes 7 to 9 correspond to the urea (0.5 M) wash fractions obtained following incubation of plasma IgG fractions with the respective recombinant antigens. The full-length, uncropped blot is provided as S1 Fig.

### 3.4 The middle-down strategy provides high specificity, enabling IGHG allele discrimination

Because of the high similarity of the protein sequences encoded by the IGHG alleles, their discrimination requires higher coverage to improve the analytical specificity. In a preliminary step, we implemented a middle-down strategy [16], and demonstrated IGHG allele molecular characterization of intact Fc/2 fragments (∼211 aa) by nano-LC-MS/MS of polyclonal total IgG. For this purpose, the total plasma IgG of the HVDtot donor were enriched on protein G, deglycosylated (PNGase F), cleaved with IdeS, and reduced with TCEP before nano-LC-MS/MS on a Q Exactive with HCD fragmentation (S2 Fig). Up to 50% sequence coverage per allele was achieved, enabling discrimination of variants with one-to-three-point mutations in CH2–CH3-CHS. Two challenges remained: applying the approach to low-abundance Ag-specific IgG, and resolving complexity from co-eluted IgG fragments from two individuals. Co-elution of near-isobaric Fc/2 variants on the PepMap300 C4 nano-LC column was a major bottleneck. The Poroshell 300SB-C3 (300 Å) improved the Fc/2 spreading along the retention time axis with reproducible elution profiles. However, the transition to UHPLC scale (80 µL/min vs. 180 nL/min) resulted in a ∼500-fold increase in flow rate, accompanied by a corresponding loss of Electrospray Ionization sensitivity, which was incompatible with low-abundance *T. gondii*-specific fractions.

### 3.5 Mass spectrometric characterization of IGHG protein alleles in the *T. gondii*-specific IgG samples isolated from a seronegative and seropositive plasma sample pair

The middle-down workflow was applied to *T. gondii*-specific IgG fractions from HVD+ and HVD− donors across four purification campaigns (2023–2025). According to the DNA sequencing of *IGHG* genes from the two donors, HVD+ carries *IGHG1*\*03, *IGHG1*\*07, *IGHG3*\*11/*12 and *IGHG3*\*22; HVD− is homozygous for *IGHG1*\*02/*08/*09/*14 and carries *IGHG3*\*14 and *IGHG3*\*16. *IGHG2* and *IGHG4* allelic status of both donors remains to be confirmed (S3 Table). After Ag affinity purification, Fc/2 fragments were fractionated by sequential IgG3/IgG1 affinity, IdeS proteolysis, TCEP reduction, and FcXP resin enrichment, yielding four fractions (FcX3−/+, FcX1−/+). Deconvoluted mass spectra confirmed correct purification, but signal intensity in the Ag-specific fractions (FcX3+, FcX1+) was substantially lower than in non-specific fractions, reflecting the low abundance of Ag-specific IgG (∼0.05–0.5% of total plasma IgG). The IgG3-specific fraction (FcX3+) was consistently at or below detection threshold. Despite multiple optimization attempts, no IGHG allele could be unambiguously assigned from intact Fc/2 mass data, prompting a transition to bottom-up proteomics of the remaining Fc/2 capture material. *T. gondii*-specific pooled IgG fractions from HVD+ and HVD− were processed for bottom-up proteomics analysis. *In silico* multi-enzyme digestion of the full IMGT database predicted all theoretically observable peptides spanning polymorphic positions. Bottom-up analysis achieved 77.8% IGHG1 sequence coverage, demonstrating sensitivity under challenging conditions. Peptide identification was performed in both DDA and DIA modes, with cross-validation between strategies.

### 3.6 No IGHG3 peptide unambiguously identified an allele

The peptide EEQ**F**NSTFR, bearing the Y296F substitution characteristic of IGHG3*11 (expected in HVD+), was detected in DIA mode but is also present in IGHG2*17 and IGHG2*02, precluding IGHG3 specificity in a polyclonal mixture (S3 Fig). Its DIA-NN score was also below the validation threshold. IGHG3 allele characterization was not pursued.

### 3.7 IGHG1 allele-specific unique signature was detected

The CH3 peptide WQQGNVFSCSVMHE**G**LHNHYTQK, bearing the A431G substitution exclusive to IGHG1*07 across the entire IGHG1 repertoire, was validated by MS/MS sequencing. The fragmentation profiles in both DDA (Xcorr 5.84, 3+) and DIA (Xcorr 7.62, 5+), provided unambiguous assignations (Fig 4 and S4 Fig). It was detected exclusively in the HVD+ fraction, consistent with DNA sequencing data, and absent in HVD−, confirming detection specificity. The closely related peptide WQQGNVFSCSVMHE**A**LHNHYTQK (G314A) was detected by DDA (Xcorr 7.72) and in DIA, but DIA-NN fragment ion scores contained >50% zero-intensity values, preventing formal DIA validation. Compatible with IGHG1*03 (present in HVD+), it cannot serve as an allele-specific signature as it is shared by numerous other IGHG1 alleles and all IGHG2 alleles, and is therefore expected in both fractions. A third peptide, WQQGN**I**FSCSVMHEALHN**R**FTQK (two substitutions: V422I, H435R), showed low confidence (Xcorr 2.08, 1+), is also shared with IGHG3, and was attributed to IGHG3*11 in HVD+. It was not retained for allele assignment.

**Fig 4.**
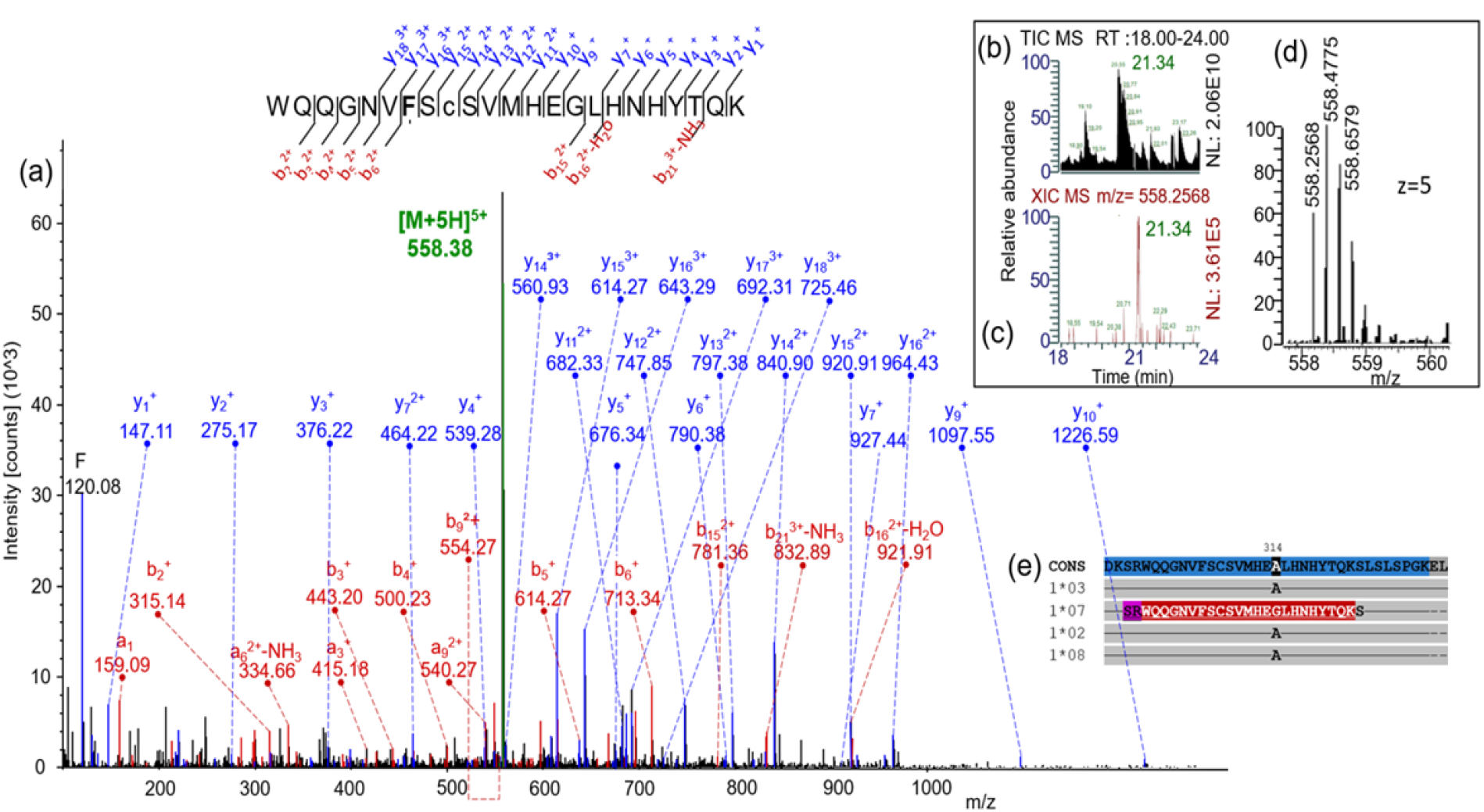
Mass spectrometric identification of an IGHG1 allele-specific peptide in *T. gondii*-specific purified IgG. Nano-LC-MS/MS features specific to the detection of the immunoglobulin heavy chain peptide WQQGNVFSCSVMHEGLHNHYTQK (IGHG1*07, charge state 3+, Xcorr = 5.84), detected exclusively in the *T. gondii* seropositive sample (HVD+). The substitution of alanine (A) by glycine at this position 314 (G, bold) is exclusive to the IGHG1*07 allele across the entire IGHG1 allelic repertoire. This peptide was not detected in the seronegative sample (HVD-). **(a)** Annotated CID Astral MS/MS spectrum generated in DDA mode for the precursor ion m/z 558.2568 (z = 5), annotated using SequestHT (tolerance 0.05 Da), showing the carbamidomethylated peptide sequence. Selected a-, b- and y-ion series are annotated. **(b)** Total ion chromatogram (TIC) over the RT window 18–24 min. **(c)** Extracted ion chromatogram (XIC) of the precursor ion m/z 558.2568, highlighting the retention time of the detected peptide. **(d)** Full scan MS profile with the isotopic distribution of the precursor ion (z = 5). **(e)** Partial amino acid sequence alignment of IGHG1 alleles in the CH3 domain, centered on polymorphic position 314. Dashes indicate amino acid identity with the consensus sequence. Alleles carrying the A residue (IGHG1*03, *02, *08 and others) are distinguished from IGHG1*07 (G residue) at this single position. The proteolytic peptide spanning this polymorphic site is underlined.

Taken together, bottom-up MS of *T. gondii*-specific IgGs purified from a mixture of a seropositive and seronegative plasma sample pair enabled unambiguous identification of the IGHG1*07-defining peptide exclusively in the HVD+ donor, in full concordance with molecular reference data. This constitutes a proof of concept for mass spectrometric detection of individual IGHG protein alleles in Ag-specific polyclonal IgG fractions. Detailed sequence assignment is provided in S4 Table.

## 4. DISCUSSION

### 4.1 Isolation of highly specific plasma IgG from *T. gondii*: choice of recombinant antigens, diagnostic value, limitations

Among the three recombinant *T. gondii* Ags produced in this study, *Tg*SAG1 proved to be the most reliable for the diagnosis of toxoplasmosis, as previously demonstrated [31]. Given that *Tg*AMA1 also exhibited good sensitivity and specificity, it could effectively complement *Tg*SAG1. The recombinant *Tg*AMA1 Ag demonstrated promising potential for the early diagnosis of *T. gondii*, particularly in its the full-length protein form (as used in our study), which shows stronger interaction with IgG (99.4%) and IgM (80.0%) than isolated N-terminal or C-terminal fragments [21]. *Tg*GRA7 proved to be the least sensitive and least specific of the Ags studied. It is likely less reliable when used alone but useful when combined with other Ags to improve the accuracy of serological tests for diagnosing acute and chronic toxoplasmosis, as suggested by Mamaghani *et al.* (2020) [32]. Furthermore, the development of rapid diagnostic tests, such as DOT-ELISA, has shown that *Tg*GRA7 exhibits 100% sensitivity and 97.2% specificity for IgM detection when combined with *Tg*SAG1. For IgG detection, the combination of *TgS*AG1 and *Tg*GRA7 achieved 100% sensitivity, confirming the utility of *Tg*GRA7 in the rapid serological diagnosis of acute and chronic toxoplasmosis, whether in the laboratory or in the field [33]. In general, *Tg*SAG1 appears to be the best choice if a single Ag is used, whereas combining it with other Ags would likely yield a more comprehensive diagnostic strategy by reducing the proportion of false positives [34]. Among the combinations identified above, the individual IgG response against *Tg*SAG1, combined with responses against *Tg*GRA7, *Tg*AMA1, or both, demonstrated the highest values for sensitivity, specificity, PPV and NPV. Hence, it was deemed the optimal combination for accurate serological diagnosis. With our aim of developing a tool for the early detection of toxoplasmosis, combining *Tg*GRA7 with the key antigen *Tg*SAG1 could enable to capture the IgG kinetics across the various phases of human *T. gondii* infection and optimize the detection of parasite-specific IgGs. Furthermore, the addition of *Tg*AMA1 proves highly useful in compensating for the inconsistency of *Tg*GRA7.

We therefore purified specific IgG directed against the 3 recombinant Ags. We initially chose to purify *T. gondii*-specific IgGs separately, using columns coated with a single Ag at a time to avoid any antigenic competition or steric hindrance that might occur with columns coated with multiple Ags. Indeed, antigenic competition can arise when multiple Ags are in contact, leading to the preferential selection of IgGs directed against specific Ags. For example, the study of Shnawa *et al.* (2023) [35] demonstrated that the simultaneous presence of *E. coli* and *P. aeruginosa* Ags in an isolation column resulted in lower retention of specific IgGs compared to the retention observed when the Ags specific to each pathogen were presented separately. The IgG fraction purified from our toxoplasmosis seropositive (HVD+) plasma contained a higher proportion of anti-*Tg*SAG1 IgGs compared to IgGs directed against the other two Ags, which were present only at residual levels, particularly in the case of *Tg*GRA7. This result is consistent with the fact that a former *T. gondii*-seropositive individual maintains a robust and long-lasting Ab response against *Tg*SAG1 [31]. However, a method combining all three Ags would be more advantageous in terms of processing time and purification yield, especially if the available plasma volume requires an iterative method necessitating the reuse of the same plasma for each Ag column. We plan to reach this final stage by combining our three recombinant Ags into a single chimeric Ag to optimize the purification protocol. Thus, the use of a single chimeric Ag could improve diagnostic performance for the early detection of *T. gondii* infection [36].

Sensitivity and specificity are generally determined against a single gold-standard test; however, in our study, serological results for toxoplasmosis were obtained using various commercial diagnostic tests (Platelia™ Toxo IgG; recomWell Toxoplasma IgG; VIDAS® TOXO IgG II) depending on the population groups included. The “negative” and “positive” serological results from these tests were used to evaluate the diagnostic performance of our three recombinant *T. gondii* Ags. The Ags underlying these tests, as well as the technical conditions of their realization, were not considered. That said, potential heterogeneity in IgG levels among the commercial tests used may have contributed to enhancing the SS, SP, NPV, and PPV values obtained from these pooled results, particularly for *Tg*SAG1.

### 4.2 Characterizing *T. gondii*-specific IGHG alleles in low-volume samples paves the way for clinical proteomic applications

The unique challenge of this study -which made it far more complex than the analysis of therapeutic monoclonal Abs typically studied-laid in characterizing the allelic composition of a minor fraction of *T. gondii-*specific IgGs within a mixture of polyclonal Abs from two individuals, all while using a sample volume limited to a few microliters of plasma to ensure compatibility with clinical monitoring of newborns. From a proteomics perspective, we pursued two objectives: (1) to validate the bottom-up proteomics approach for characterizing IGHG alleles in low-abundance, Ag-specific IgG fractions, and (2) to assess whether DNA-based molecular allele assignments could be recapitulated at the protein level in a mixed-donor context. This work highlights the critical role of yield constraints when combining middle-down and bottom-up strategies.

Our foundational work [16] established the first proof-of-concept for discriminating IGHG alleles at the intact Fc/2 fragment level using LC-MS/MS on total polyclonal IgGs. Using HCD fragmentation of deglycosylated Fc/2 fragments (∼211 aa, ∼24 kDa) on a Q Exactive mass spectrometer, we achieved sequence coverage of up to 50% per allele, which proved sufficient to distinguish single amino acid substitutions. IGHG1 alleles were characterized most consistently, reflecting the predominance of IgG1 in serum (∼65–70%). However, two prerequisites for clinical application have emerged: working with ultra-low-abundance Ag-specific IgGs and resolving the complexity of samples containing IGHG alleles from two individuals (with up to 16 alleles theoretically coexisting for two unrelated individuals, or 12 alleles in the case of a mother and her newborn).

This study directly addressed both challenges. *T. gondii*-specific IgG account for only 0.05–0.5% of total plasma IgG (mean total serum IgG level: ∼10 mg/mL; anti-*Tg*SAG1 IgG level in seropositive individuals: 5–50 μg/mL, mean: 16 μg/mL; comparable responses for anti-T*g*AMA1 and anti-*Tg*GRA7 Abs). This extremely low proportion, several orders of magnitude below the 200-ng detection threshold of our proof-of-concept study, posed a major limitation for LC-MS. Despite multiple attempts under various conditions, middle-down proteomics failed to yield allele-specific spectra. Although current high-resolution instruments are 1–2 orders of magnitude more sensitive than the original Q Exactive model, and affinity enrichment could be optimized, neither factor would fully compensate for the sensitivity loss resulting from the switch from nano- to micro-LC. Miniaturizing the LC separation could ultimately solve this problem, but it requires robust, reproducible nano-LC columns with dedicated large-pore stationary phases, which are not yet commercially available.

Our results established the feasibility of detecting disease-specific IGHG alleles in purified IgGs isolated from a pooled plasma sample – comprising material from one seronegative and one seropositive donor for toxoplasmosis - whose IGHG protein alleles had been previously identified via DNA molecular analysis. This work enabled the detection of amino acid polymorphisms in the Fc/2 fragment of the *T. gondii*-specific IgG and the identification of the IGHG1*07 allele exclusively in the seropositive sample, consistent with DNA sequencing data. However, further optimization is required before considering clinical application of this approach. Middle-down proteomics offers, in principle, analytical superiority: it remains the most specific strategy for distinguishing IGHG alleles as it targets intact Fc/2 fragments, thus avoiding ambiguities associated with shorter tryptic peptide sequences common to multiple alleles and subclasses. Chromatographic separation of quasi-isobaric Fc/2 variants poses the primary analytical challenge. Although some stationary phases offer the required selectivity, they are not yet available in the nanoscale format (internal diameter ≤ 320 μm) required to achieve the sensitivity characteristic of nano-LC. Consequently, the in-house fabrication of packed capillary columns with wide-pore C3 or Polystyrene-divinylbenzene stationary phases, in a format robust and reproducible enough for routine nano-LC use, represents a key technical objective for advancing the middle-down strategy in this study. Integrating this approach into the diagnostic workflow should be the next logical step, leveraging the sensitivity gains offered by the current generation of instruments.

Sensitivity remains the primary bottleneck: *T. gondii*-specific IgG are highly diluted relative to total IgG, and neonatal samples impose even stricter volume constraints. The use of multi-Ag affinity columns combining *Tg*SAG1, *Tg*AMA1 and *Tg*GRA7 in a single step would increase the yield of specific IgGs and reduce processing time, a design already considered in the purification strategy. Furthermore, DDA and DIA approaches are complementary: DDA provides robust spectral peptide matches, whereas DIA offers broader and more reproducible peptide coverage. This highlights the need to integrate targeted DDA-based confirmation into future developments. Finally, it will be necessary to extend the search for allele-specific peptides to the IGHG2 and IGHG4 subclasses, as well as to other donors with fully characterized genetic profiles (DNA sequences), in order to evaluate the generalizability of this proteotypic strategy across the full diversity of human IGHG alleles. The resulting development priorities, and the stage they define in the maturation of the approach, are summarized in the Conclusion.

## 5. CONCLUSION

The present results demonstrate, as a proof of concept, that IGHG alleles can be detected within disease-specific IgG purified from a mixture of plasma from two distinct donors. The objective is a postnatal test able to attribute *T. gondii*-specific IgG to the newborn rather than to its mother, from a few microliters of blood and within the first weeks of life, at a stage where conventional serology cannot discriminate between the two origins. The step established here is the central link of that chain: parasite-specific IgG can be isolated with sufficient purity on recombinant antigens, and the resulting polyclonal fraction, although it represents less than 0.5% of total plasma IgG, still carries an allele-specific signature that bottom-up proteomics assigns unambiguously, in agreement with molecular data.

Middle-down proteomics nevertheless remains the ideal analytical strategy, as it reads the polymorphic positions within a single intact Fc/2 polypeptide and thereby avoids the allele and subclass ambiguities inherent in short tryptic peptides. As detailed in the Discussion, its deployment is held back by two coupled constraints, namely the chromatographic resolution of quasi-isobaric Fc/2 variants and the sensitivity loss imposed by the chromatographic scale at which that resolution is currently achievable; lifting them jointly, through dedicated nano-scale wide-pore columns, is the principal development ahead. The complementary steps identified above, from the yield of the immunopurification to the coverage of the IGHG2 and IGHG4 subclasses and the extension to further sequenced donors, and then to true mother-newborn pairs, define the remaining path to clinical evaluation.

In summary, this study complements current research focused on serum IgGs for therapeutic purposes [37,38]. It paves the way for an alternative diagnostic method capable of improving the likelihood of early detection of congenital toxoplasmosis and, more broadly, clinical outcomes of infants with congenital diseases.

## Data Availability

All data produced in the present study are available upon reasonable request to the authors

## AUTHOR CONTRIBUTIONS

Study concept and design: MD, CD, FMN, JV; obtained funding: MD, CD, FMN, JV; acquisition of data: MS, ED, MD, AE, ZK, SD, NS, CD, FMN; analysis of data: MS, ED, AE, CD, JV; drafting of the manuscript: MS, ED, MD, AE, CD, FMN, JV; critical revision of the manuscript: MS, MD, ZK, SD, NS, CD, FMN, JV; study supervision: MD, CD, FMN, JV; administration: FMN, JV. All authors gave their final approval and agreed to be accountable for all aspects of the work.

## ACKNOWLEDGEMENTS

We thank the women involved in the studies in Benin (PlasDCty and CoaLa projects) and in France (TOXODIAG project) for their valuable participation. We are grateful to the field medical staff from the various maternity wards for their valuable contribution to the project: midwives, nurses, and biological technicians. The TOXODIAG sponsor was Assistance Publique – Hôpitaux de Paris (Délégation à la Recherche Clinique et à l’Innovation).

## DATA AVAILABILITY STATEMENT

The data supporting the findings of this study are available from the corresponding authors (JV and MS) upon reasonable request.

## Abbreviations

Ab: Antibody
Ag: Antigen
AUC: Area Under the Curve
BSA: Bovine Serum Albumin
CH: Constant domain of the immunoglobulin heavy chain
CT: Congenital Toxoplasmosis
DDA: Data-Dependent Acquisition
DIA: Data-Independent Acquisition
DNA: Deoxyribonucleic acid
DTT: Dithiothreitol
ELISA: Enzyme-Linked ImmunoSorbent Assay
Fc: Fragment crystallizable region
HCD: Higher-energy Collisional Dissociation
HRP: Horseradish peroxidase
Ig: Immunoglobulin
IGHG: ImmunoGlobulin Heavy G chain
IMGT®: the international ImMunoGeneTics information system®
LC: Liquid Chromatography
MS: Mass spectrometry
PBS: Phosphate-Buffered Saline
PNV: Negative Predicted Value
PPV: Positive Predicted Value
ROC: Receiver Operating Characteristic
SP: Specificity
SS: Sensitivity
TCEP: Tris(2-carboxyethyl) phosphine hydrochloride
*Tg*AMA1: *Toxoplasma gondii* Apical Membrane Antigen 1
*Tg*GRA7: *Toxoplasma gondii* Dense Granule Protein 7
*Tg*SAG1: *Toxoplasma gondii* Surface Antigen 1
UHPLC: Ultra High-Performance Liquid Chromatography

## SUPPORTING INFORMATION

**S1 Fig.** Full-length, uncropped and non-adjusted Western blot corresponding to Fig 3.

**S2 Fig.** LC-MS characterization of the Fc/2 Fragments of total IgG1 and IgG3 from the HVDtot donor.

**S3 Fig.** Sequence alignment of identified IGHG1 and IGHG3 peptides against the corresponding allelic sequences.

**S4 Fig.** DIA MS/MS spectrum confirming the detection of the IGHG1*07 peptide.

**S1 Table.** Amino acid variations in the CH1, CH2 and CH3-CHS constant domains, and in the Hinge regions of IGHG.

**S2 Table.** Equimolar distribution of recombinant *T. gondii* antigens used for the immunopurification columns.

**S3 Table**. Molecular determination of *IGHG* alleles in the three donors: HVDtot, HVD+, and HVD-.

**S4 Table.** Mass spectrometric identification of IGHG peptides by DDA and DIA, peptide sequence, compatible allele(s), DDA Xcorr score, DIA-NN score, charge state, detection in HVD+ and HVD-samples, and validation status.

**S1 File.** Detailed protocols for the production of recombinant *T. gondii* antigens and the characterization of IGHG alleles by mass spectrometry.

