## Supplementary material for "Isolation of *Toxoplasma gondii*-specific IgG and mass spectrometric characterization of individual IgGs for early diagnosis of toxoplasmosis": S1_Fig

### **S1 Fig. Full-length, uncropped and non-adjusted Western blot corresponding to Fig 3.**

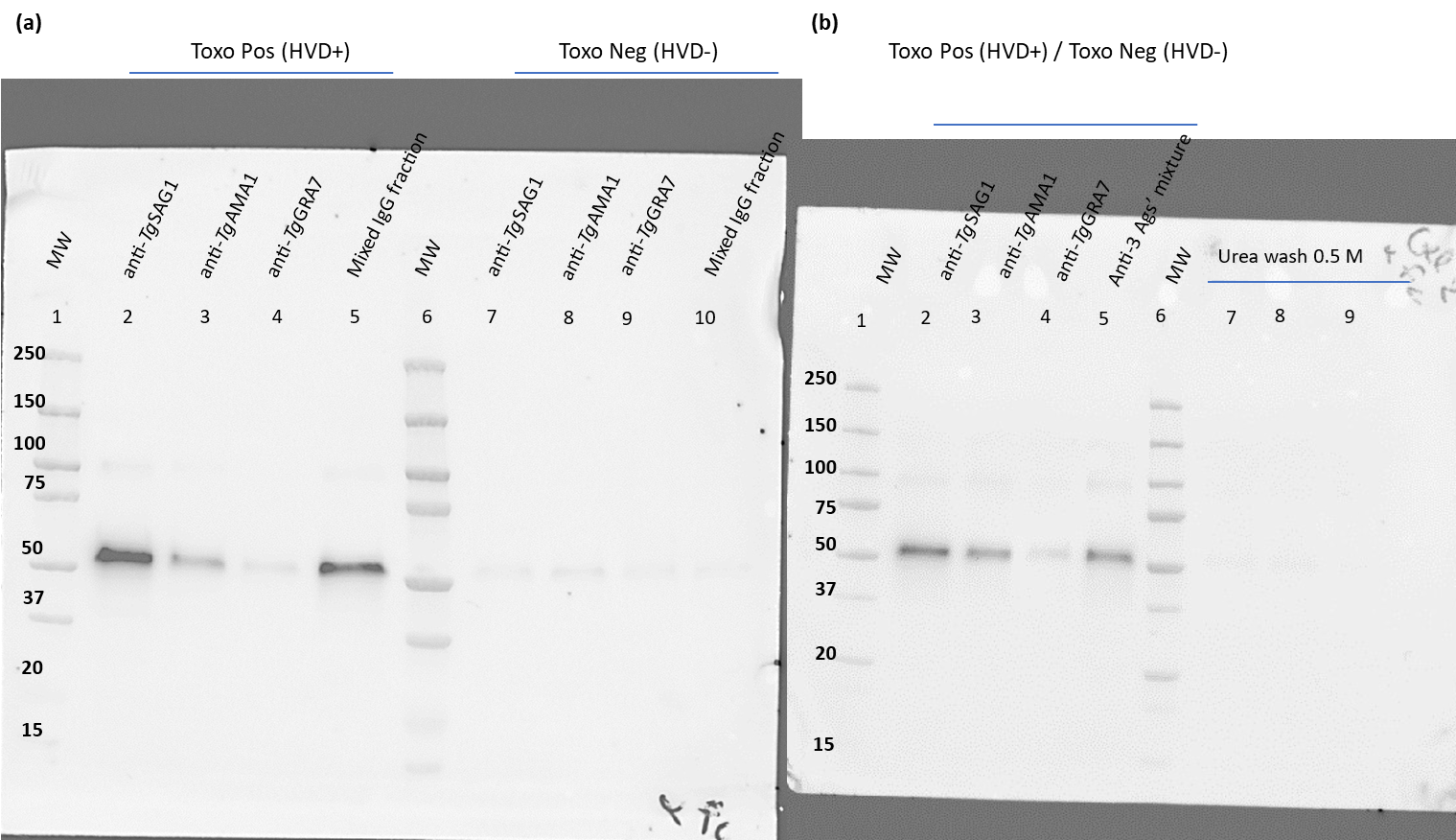

Recognition of the produced *Tg*SAG1, *Tg*AMA1 and *Tg*GRA7 recombinant antigens by IgG purified from *T. gondii* seronegative and seropositive samples (Unprocessed Western blot):

**(a)** Toxo Pos (HVD+): plasma sample seropositive for toxoplasmosis; Toxo Neg (HVD-): plasma sample toxoplasmosis seronegative. Lanes 2 to 4 correspond to the specific IgG fractions directed against the recombinant antigens *Tg*SAG1, *Tg*AMA1 and *Tg*GRA7, respectively, from HVD+. Lanes 7 to 9 correspond to the antigen-specific IgG fractions from HVD-. Lane 5: the mixed IgG fraction corresponds to a mixture of the IgG fractions directed against each antigen, in equal volume for HVD+ (lanes 2 to 4) and HVD- (fractions 7 to 9).

**(b)** Toxo Pos (HVD+) / Toxo Neg (HVD-): an equal-volume mixture of plasma samples seropositive and seronegative for toxoplasmosis, respectively. Lanes 2 to 4 show the cumulative IgG responses against the recombinant antigens *Tg*SAG1, *Tg*AMA1, and *Tg*GRA7, respectively. Lane 5 shows the cumulative IgG response against a mixture of the three recombinant antigens combined in equal volumes. Lanes 7 to 9 correspond to the urea (0.5 M) wash fractions obtained following incubation of plasma IgG fractions with the respective recombinant antigens.
