## Supplementary material for "Isolation of *Toxoplasma gondii*-specific IgG and mass spectrometric characterization of individual IgGs for early diagnosis of toxoplasmosis": S2_Fig

### S2 Fig. LC-MS characterization of the Fc/2 Fragments of total IgG1 and IgG3 from the HVDtot donor.

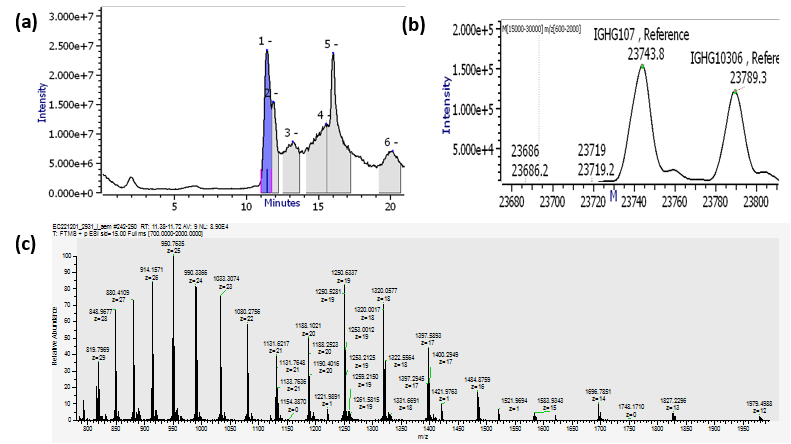

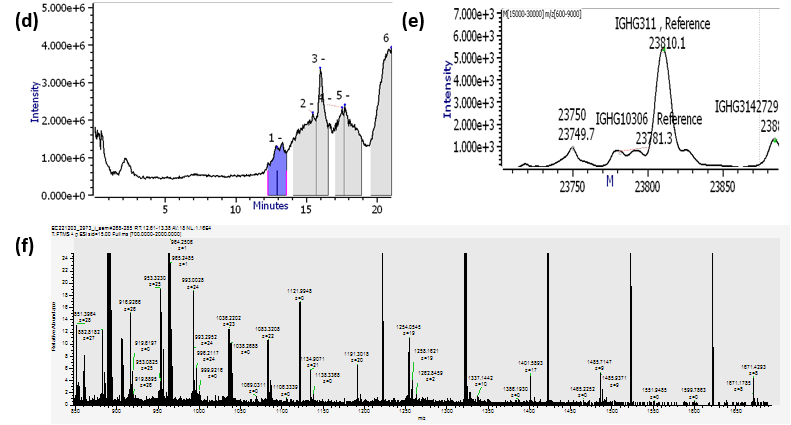

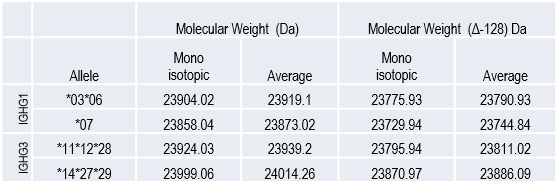

Characterization of IgG1 and IgG3 Fc/2 fragments by LC-MS and detection following enrichment of total plasma IgG from the HVDtot donor: IgG1 Fc/2: **(a)** LC chromatogram **(b)** deconvoluted mass profile, **(c)** raw MS charge state envelope; IgG3 Fc/2: **(d)** LC chromatogram, **(e)** deconvoluted mass profile, **(f)** raw MS charge state envelope.

The table in the lower part lists theoretical IGHG1 and IGHG3 allele masses, including correction for C-terminal lysine loss (–128 Da) and associated processing variants. The Fc/2 fragments of human IgG1 and IgG3 were enriched from a polyclonal model serum after molecular characterization of the allelic composition using subclass-specific nanobodies (CaptureSelect™, ThermoFisher Scientific) prior LC-MS analysis as described in the material and method section for IGHG allele characterization. For IgG1 Fc/2 experiment, 20µL of serum (containing approximately 200 µg of total IgG, including 130 µg of IgG1) were incubated with 20 µL of CaptureSelect™ IgG1-specific nanobodies (binding capacity >160 µg). For IgG3 Fc/2 experiment, 30µL of serum (containing approximately 300 µg of total IgG, including 21 µg of IgG3) were incubated with 20 µL of CaptureSelect™ IgG3-specific nanobodies (binding capacity >120 µg). After immunocapture, bound IgG were eluted with 40 µL of 1% formic acid and immediately neutralized with 40 µL of 1.5 M Tris-HCl (pH 8.8).

Data interpretation was performed using Intact Mass Protein Metrics and Freestyle deconvolution software.
