## Supplementary material for "Isolation of *Toxoplasma gondii*-specific IgG and mass spectrometric characterization of individual IgGs for early diagnosis of toxoplasmosis": S3_Fig

### **S3 Fig. Sequence alignment of identified IGHG1 and IGHG3 peptides against the corresponding allelic sequences.**

**A	Alignment IGHG1 and identified sequences**

Identified with Trypsin+LysC/LysC/both

1———————————————————————————————————————CH1——————————————————————————————————70

|  | Series 1 | Series 2 | Series 3 | Series 4 | Series 5 | Series 6 | Series 7 | Series 8 | Series 9 | Series 10 | Series 11 | Series 12 | Series 13 | Series 14 | Series 15 | Series 16 | Series 17 | Series 18 | Series 19 | Series 20 |
| --- | --- | --- | --- | --- | --- | --- | --- | --- | --- | --- | --- | --- | --- | --- | --- | --- | --- | --- | --- | --- |
| Category 1 | 4 | 4 | 4 | 4 | 4 | 4 | 4 | 4 | 4 | 4 | 4 | 4 | 4 | 4 | 4 | 4 | 4 | 4 | 4 | 4 |

```
CONS1  ASTKGPSVFP LAPSSKSTSG GTAALGCLVK DYFPEPVTVS WNSGALTSGV HTFPAVLQSS GLYSLSSVVT
1*03   ————GPSVFP LAPSSKSTSG GTAALGCLVK —————————— —————————— —————————— ——————————
1*07   ————GPSVFP LAPSSKSTSG GTAALGCLVK —————————— —————————— —————————— ——————————
1*02   ————GPSVFP LAPSSKSTSG GTAALGCLVK —————————— —————————— —————————— ——————————
1*08   ————GPSVFP LAPSSKSTSG GTAALGCLVK —————————— —————————— —————————— ——————————
```

71——————————————CH1————————————————————|——————Hinge—————|————————CH2——————————140

|  | Series 1 | Series 2 | Series 3 | Series 4 | Series 5 | Series 6 | Series 7 | Series 8 | Series 9 | Series 10 | Series 11 | Series 12 | Series 13 | Series 14 | Series 15 | Series 16 | Series 17 | Series 18 | Series 19 | Series 20 |
| --- | --- | --- | --- | --- | --- | --- | --- | --- | --- | --- | --- | --- | --- | --- | --- | --- | --- | --- | --- | --- |
| Category 1 | 4 | 4 | 4 | 4 | 4 | 4 | 4 | 4 | 4 | 4 | 4 | 4 | 4 | 4 | 4 | 4 | 4 | 4 | 4 | 4 |

```
CONS1  VPSSSLGTQT YICNVNHKPS NTKVDKRVEP KSCDKTHTCP PCPAPELLGG PSVFLFPPKP KDTLMISRTP
1*03   —————————— —————————— ——————RVEP KSCDKTHTCP PCPAPELLGG PSVFLFPPKP KDTLMISRTP
1*07   —————————— —————————— ——————RVEP KSCDKTHTCP PCPAPELLGG PSVFLFPPKP KDTLMISRTP
1*02   —————————— —————————— ——————K——— —SCDKTHTCP PCPAPELLGG PSVFLFPPKP KDTLMISRTP
1*08   —————————— —————————— ——————RVEP KSCDKTHTCP PCPAPELLGG PSVFLFPPKP KDTLMISRTP
```

141—————————————————————————————————————CH2————————————————————————————————————210

|  | Series 1 | Series 2 | Series 3 | Series 4 | Series 5 | Series 6 | Series 7 | Series 8 | Series 9 | Series 10 | Series 11 | Series 12 | Series 13 | Series 14 | Series 15 | Series 16 | Series 17 | Series 18 | Series 19 | Series 20 |
| --- | --- | --- | --- | --- | --- | --- | --- | --- | --- | --- | --- | --- | --- | --- | --- | --- | --- | --- | --- | --- |
| Category 1 | 4 | 4 | 4 | 4 | 4 | 4 | 4 | 4 | 4 | 4 | 4 | 4 | 4 | 4 | 4 | 4 | 4 | 4 | 4 | 4 |

```
CONS1  EVTCVVVDVS HEDPEVKFNW YVDGVEVHNA KTKPREEQYN STYRVVSVLT VLHQDWLNGK EYKCKVSNKA
1*03   EVTCVVVDVS HEDPEVKFNW YVDGVEVHNA K————EEQYN STYRVVSVLT VLHQDWLNGK EYKCKVSNKA
1*07   EVTCVVVDVS HEDPEVKFNW YVDGVEVHNA K————EEQYN STYRVVSVLT VLHQDWLNGK EYKCKVSNKA
1*02   EVTCVVVDVS HEDPEVKFNW YVDGVEVHNA K————EEQYN STYRVVSVLT VLHQDWLNGK EYKCKVSNKA
1*08   EVTCVVVDVS HEDPEVKFNW YVDGVEVHNA K————EEQYN STYRVVSVLT VLHQDWLNGK EYKCKVSNKA
```

211————————————————————————————————————CH2———————————————————————————————|—CH3—280

|  | Series 1 | Series 2 | Series 3 | Series 4 | Series 5 | Series 6 | Series 7 | Series 8 | Series 9 | Series 10 | Series 11 | Series 12 | Series 13 | Series 14 | Series 15 | Series 16 | Series 17 | Series 18 | Series 19 | Series 20 |
| --- | --- | --- | --- | --- | --- | --- | --- | --- | --- | --- | --- | --- | --- | --- | --- | --- | --- | --- | --- | --- |
| Category 1 | 4 | 4 | 4 | 4 | 4 | 4 | 4 | 4 | 4 | 4 | 0 | 4 | 4 | 4 | 4 | 4 | 4 | 4 | 4 | 4 |

```
CONS1  LPAPIEKTIS KAKGQPREPQ VYTLPPSRDE LTKNQVSLTC LVKGFYPSDI AVEWESNGQP ENNYKTTPPV
1*03   LPAPIEK——— ———————EPQ VYTLPPSRE— M——NQVSLTC LVK——————— —————————— —————TTPPV
1*07   LPAPIEK——— ———————EPQ VYTLPPSRD— L——NQVSLTC LVK——————— —————————— —————TTPPV
1*02   LPAPIEK——— ———————EPQ VYTLPPSRD— L——NQVSLTC LVK——————— —————————— —————TTPPV
1*08   LPAPIEK——— ———————EPQ VYTLPPSRD— L——NQVSLTC LVK——————— —————————— —————TTPPV
```

281——————————————————————CH3—————————————————————————————|—————————————————————350

|  | Series 1 | Series 2 | Series 3 | Series 4 | Series 5 | Series 6 | Series 7 | Series 8 | Series 9 | Series 10 | Series 11 | Series 12 | Series 13 | Series 14 | Series 15 | Series 16 | Series 17 | Series 18 | Series 19 | Series 20 |
| --- | --- | --- | --- | --- | --- | --- | --- | --- | --- | --- | --- | --- | --- | --- | --- | --- | --- | --- | --- | --- |
| Category 1 | 4 | 4 | 4 | 4 | 4 | 4 | 4 | 4 | 4 | 4 | 0 | 4 | 4 | 4 | 4 | 4 | 4 | 4 | 4 | 4 |

```
CONS1  LDSDGSFFLY SKLTVDKSRW QQGNVFSCSV MHEALHNHYT QKSLSLSPGK ELQLEESCAE AQDGELDGLW
1*03   LDSDGSFFLY SK———————— —————————— ———A—————— —————————— —————————— ——————————
1*07   LDSDGSFFLY SK—————SRW QQGNVFSCSV MHEGLHNHYT QK———————— ---------- ----------
1*02   LDSDGSFFLY SK———————— —————————— ———A—————— —————————— ---------- ----------
1*08   LDSDGSFFLY SK———————— —————————— ———A—————— —————————— ---------- ----------
```

351————————————————————————————————————————————————————————401

|  | Series 1 | Series 2 | Series 3 | Series 4 | Series 5 | Series 6 | Series 7 | Series 8 | Series 9 | Series 10 | Series 11 | Series 12 | Series 13 | Series 14 | Series 15 | Series 16 | Series 17 | Series 18 | Series 19 | Series 20 |
| --- | --- | --- | --- | --- | --- | --- | --- | --- | --- | --- | --- | --- | --- | --- | --- | --- | --- | --- | --- | --- |
| Category 1 | 1 | 1 | 1 | 1 | 1 | 1 | 1 | 1 | 1 | 1 | 0 | 1 | 1 | 1 | 1 | 1 | 1 | 1 | 1 | 1 |

```
CONS1  TTITIFITLF LLSVCYSATV TFFKVKWIFS SVVDLKQTII PDYRNMIGQG A
1*03   —————————— —————————— —————————— ——————QTII PDYRNMIGQG A
1*07   ---------- ---------- ---------- ---------- ---------- -
1*02   ---------- ---------- ---------- ---------- ---------- -
```

1*08	---------- ---------- ---------- ---------- ---------- -

**B	Alignment IGHG3 and identified sequences**

Identified with Trypsin+LysC/LysC/both

|  | Series 1 | Series 2 | Series 3 | Series 4 | Series 5 | Series 6 | Series 7 | Series 8 | Series 9 | Series 10 | Series 11 | Series 12 | Series 13 | Series 14 | Series 15 | Series 16 | Series 17 | Series 18 | Series 19 | Series 20 |
| --- | --- | --- | --- | --- | --- | --- | --- | --- | --- | --- | --- | --- | --- | --- | --- | --- | --- | --- | --- | --- |
| Category 1 | 5 | 5 | 5 | 5 | 5 | 5 | 5 | 5 | 5 | 5 | 5 | 5 | 5 | 5 | 5 | 5 | 5 | 5 | 5 | 5 |

```
     1———————————————————————————————————————CH1———————————————————————————————————80
CONS  ASTKGPSVFP LAPCSRSTSG GTAALGCLVK DYFPEPVTVS WNSGALTSGV HTFPAVLQSS GLYSLSSVVT VPSSSLGTQT
3*11  ASTKGPSVFP LAPCSRSTSG GTAALGCLVK DYFPEPVTVS WNSGALTSGV HTFPAVLQSS GLYSLSSVVT VPSSSLGTQT
3*12  ASTKGPSVFP LAPCSRSTSG GTAALGCLVK DYFPEPVTVS WNSGALTSGV HTFPAVLQSS GLYSLSSVVT VPSSSLGTQT
3*22  ASTKGPSVFP LAPCSRSTSG GTAALGCLVK DYFPEPVTVS WNSGALTSGV HTFPAVLQSS GLYSLSSVVT VPSSSLGTQT
3*14  ASTKGPSVFP LAPCSRSTSG GTAALGCLVK DYFPEPVTVS WNSGALTSGV HTFPAVLQSS GLYSLSSVVT VPSSSLGTQT
3*16  ASTKGPSVFP LAPCSRSTSG GTAALGCLVK DYFPEPVTVS WNSGALTSGV HTFPAVLQSS GLYSLSSVVT VPSSSLGTQT
```

81————————CH1—————————|—————————————————————————————————Hinge———————————————————160

|  | Series 1 | Series 2 | Series 3 | Series 4 | Series 5 | Series 6 | Series 7 | Series 8 | Series 9 | Series 10 | Series 11 | Series 12 | Series 13 | Series 14 | Series 15 | Series 16 | Series 17 | Series 18 | Series 19 | Series 20 |
| --- | --- | --- | --- | --- | --- | --- | --- | --- | --- | --- | --- | --- | --- | --- | --- | --- | --- | --- | --- | --- |
| Category 1 | 5 | 5 | 5 | 5 | 5 | 5 | 5 | 5 | 5 | 5 | 0 | 5 | 5 | 5 | 5 | 5 | 5 | 5 | 5 | 4 |

```
CONS  YTCNVNHKPS NTKVDKRVEL KTPLGDTTHT CPRCPEPKSC DTPPPCPRCP EPKSCDTPPP CPRCPEPKSC DTPPPCPRCP
3*11  YTCNVNHKPS NTKVDKRV-L KTPLGDTTHT CPRCPEPKSC DTPPPCPRCP EPKSCDTPPP CPRCPEPKSC DTPPPCPRCP
3*12  YTCNVNHKPS NTKVDKRVEL KTPLGDTTHT CPRCPEPKSC DTPPPCPRCP EPKSCDTPPP CPR------- --------CP
3*22  YTCNVNHKPS NTKVDKRVEL KTPLGDTTHT CPRCPEPKSC DTPPPCPRCP EPKSCDTPPP CPRCPEPKSC DTPPPCPRCP
3*14  YTCNVNHKPS NTKVDKRVEL KTPLGDTTHT CPRCPEPKSC DTPPPCPRCP EPKSCDTPPP CPRCPEPKSC DTPPPCPRCP
3*16  YTCNVNHKPS NTKVDKRVEL KTPLGDTTHT CPRCPEPKSC DTPPPCPRCP EPKSCDTPPP CPRCPEPKSC DTPPPCPRCP
```

161—Hinge|——————————————————————————————————————CH2——————————————————————————————240

|  | Series 1 | Series 2 | Series 3 | Series 4 | Series 5 | Series 6 | Series 7 | Series 8 | Series 9 | Series 10 | Series 11 | Series 12 | Series 13 | Series 14 | Series 15 | Series 16 | Series 17 | Series 18 | Series 19 | Series 20 |
| --- | --- | --- | --- | --- | --- | --- | --- | --- | --- | --- | --- | --- | --- | --- | --- | --- | --- | --- | --- | --- |
| Category 1 | 5 | 5 | 5 | 5 | 5 | 5 | 5 | 5 | 5 | 5 | 0 | 5 | 5 | 5 | 5 | 5 | 5 | 5 | 5 | 5 |

```
CONS  APELLGGPSV FLFPPKPKDT LMISRTPEVT CVVVDVSHED PEVQFKWYVD GVEVHNAKTK LREEQYNSTF RVVSVLTVLH
3*11  APELLGGPSV FLFPPKPKDT LMISRTPEVT CVVVDVSHED PEVQFKWYVD GVEVHNAKTK PREEQFNSTF RVVSVLTVLH
3*12  APELLGGPSV FLFPPKPKDT LMISRTPEVT CVVVDVSHED PEVQFKWYVD GVEVHNAKTK PREEQYNSTF RVVSVLTVLH
3*22  APELLGGPSV FLFPPKPKDT LMISRTPEVT CVVVDVSHED PEVQFKWYVD GVEVHNAKTK LREEQYNSTF RVVSVLTVLH
3*14  APELLGGPSV FLFPPKPKDT LMISRTPEVT CVVVDVSHED PEVQFKWYVD GVEVHNAKTK LREEQYNSTF RVVSVLTVLH
3*16  APELLGGPSV FLFPPKPKDT LMISRTPEVT CVVVDVSHED PEVQFKWYVD GVEVHNAKTK LREEQYNSTF RVVSVLTVLH
```

241————————CH2————————————————|——————————————————————CH3—————————————————————————320

|  | Series 1 | Series 2 | Series 3 | Series 4 | Series 5 | Series 6 | Series 7 | Series 8 | Series 9 | Series 10 | Series 11 | Series 12 | Series 13 | Series 14 | Series 15 | Series 16 | Series 17 | Series 18 | Series 19 | Series 20 |
| --- | --- | --- | --- | --- | --- | --- | --- | --- | --- | --- | --- | --- | --- | --- | --- | --- | --- | --- | --- | --- |
| Category 1 | 5 | 5 | 5 | 5 | 5 | 5 | 5 | 5 | 5 | 5 | 0 | 5 | 5 | 5 | 5 | 5 | 5 | 5 | 5 | 5 |

```
CONS  QDWLNGKEYK CKVSNKALPA PIEKTISKTK GQPREPQVYT LPPSREEMTK NQVSLTCLVK GFYPSDIAVE WESNGQPENN
3*11  QDWLNGKEYK CKVSNKALPA PIEKTISKTK GQPREPQVYT LPPSREEMTK NQVSLTCLVK GFYPSDIAVE WESSGQPENN
3*12  QDWLNGKEYK CKVSNKALPA PIEKTISKTK GQPREPQVYT LPPSREEMTK NQVSLTCLVK GFYPSDIAVE WESSGQPENN
3*22  QDWLNGKEYK CKVSNKALPA PIEKTISKTK GQPREPQVYT LPPSREEMTK NQVSLTCLVK GFYPSDIAVE WESNGQPENN
3*14  QDWLNGKEYK CKVSNKALPA PIEKTISKTK GQPREPQVYT LPPSREEMTK NQVSLTCLVK GFYPSDIAVE WESNGQPENN
3*16  QDWLNGKEYK CKVSNKALPA PIEKTISKAK GQPREPQVYT LPPSREEMTK NQVSLTCLVK GFYPSDIAVE WESNGQPENN
```

321——————————————————————————CH3——————————————————————————|——————————————————————400

|  | Series 1 | Series 2 | Series 3 | Series 4 | Series 5 | Series 6 | Series 7 | Series 8 | Series 9 | Series 10 | Series 11 | Series 12 | Series 13 | Series 14 | Series 15 | Series 16 | Series 17 | Series 18 | Series 19 | Series 20 |
| --- | --- | --- | --- | --- | --- | --- | --- | --- | --- | --- | --- | --- | --- | --- | --- | --- | --- | --- | --- | --- |
| Category 1 | 5 | 5 | 5 | 5 | 5 | 5 | 5 | 5 | 5 | 5 | 0 | 5 | 5 | 5 | 5 | 5 | 5 | 5 | 5 | 5 |

```
CONS  YNTTPPMLDS DGSFFLYSKL TVDKSRWQQG NIFSCSVMHE ALHNRYTQKS LSLSPGKELQ LEESCAEAQD GELDGLWTTI
3*11  YNTTPPMLDS DGSFFLYSKL TVDKSRWQQG NIFSCSVMHE ALHNRFTQKS LSLSPGKELQ LEESCAEAQD GELDGLWTTI
3*12  YNTTPPMLDS DGSFFLYSKL TVDKSRWQQG NIFSCSVMHE ALHNRFTQKS LSLSPGK--- ---------- ----------
3*22  YNTTPPMLDS DGSFFLYSKL TVDKSRWQQG NIFSCSVMHE ALHNHYTQKS LSLSPGK--- ---------- ----------
3*14  YNTTPPMLDS DGSFFLYSKL TVDKSRWQQG NIFSCSVMHE ALHNRYTQKS LSLSPGKELQ LEESCAEAQD GELDGLWTTI
```

3*16 YN*TTPPMLDS* *DGSFFLYSK***L** **TVDKSRWQQG NIFSCSVMHE** **ALHNRYTQK**S LSLSPGK--- ---------- ----------

|  | Series 56 | Series 57 | Series 58 | Series 59 | Series 60 | Series 61 | Series 62 | Series 63 | Series 64 | Series 65 | Series 66 | Series 67 | Series 68 | Series 69 | Series 70 | Series 71 | Series 72 | Series 73 | Series 74 | Series 75 |
| --- | --- | --- | --- | --- | --- | --- | --- | --- | --- | --- | --- | --- | --- | --- | --- | --- | --- | --- | --- | --- |
| Category 1 | 2 | 2 | 2 | 2 | 2 | 2 | 2 | 2 | 2 | 2 | 0 | 2 | 2 | 2 | 2 | 2 | 2 | 2 | 2 | 2 |

```
 401————————————————————————————————————————————————448
CONS  TIFITLFLLS VCYSATVTFF KVKWIFSSVV DLKQTIIPDY RNMIGQGA
3*11  TIFITLFLLS VCYSATVTFF KVKWIFSSVV DLKQTIIPDY RNMIGQGA
3*12  ---------- ---------- ---------- ---------- --------
3*22  ---------- ---------- ---------- ---------- --------
3*14  TIFITLFLLS VCYSATVTFF KVKWIFSSVV DLKQTIIPDY RNMIGQGA
3*16  ---------- ---------- ---------- ---------- --------
```

**C	Graphical view of the alignment IGHG3 and identified sequences**

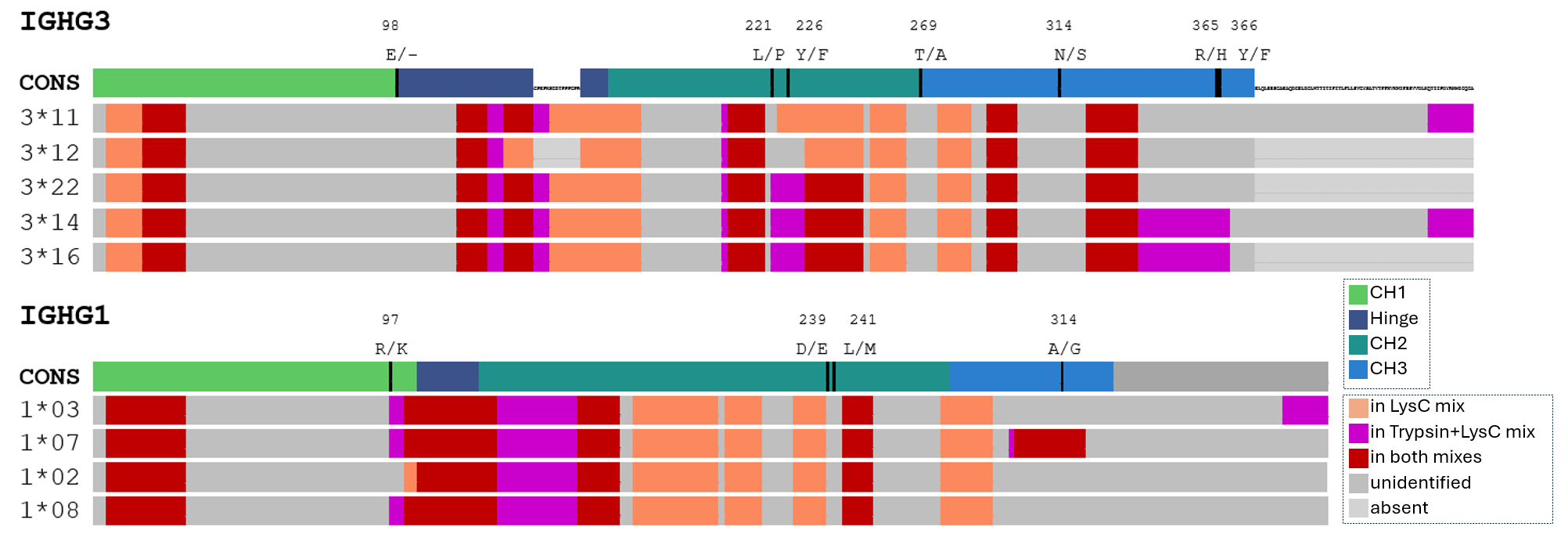

Mixture of *T. gondii*-specific IgG fractions from HVD+ and HVD- donors: sequence alignment of identified IGHG1 and IGHG3 peptides against the corresponding allelic sequences. **(A)** Alignment of peptide sequences identified by bottom-up mass spectrometry (Trypsin+LysC, LysC, or both) against the IGHG1 consensus sequence (CONS1) and four alleles (IGHG1*03, *07, *02 and *08). Domain boundaries are indicated above the alignment (CH1, Hinge, CH2, CH3). Dashes (—) indicate sequence identity with the consensus. Hyphens (–) indicate sequence not covered by any identified peptide for the corresponding allele. Bold residues in the consensus sequence denote positions where sequence variation occurs across IGHG1 alleles. The G/A substitution at position 314 (CH3 domain), diagnostic for IGHG1*07 (G) versus all other alleles (A), is highlighted. **(B)** Alignment of peptide sequences identified by bottom-up mass spectrometry (Trypsin+LysC, LysC, or both) against the IGHG3 consensus sequence (CONS) and five alleles (IGHG3*11, *12, *22, *14 and *16). Domain boundaries are indicated above the alignment (CH1, Hinge, CH2, CH3). The extended hinge region characteristic of IGHG3, with its multiple CPRCPEPKSCDTPPP repeat units, is annotated. Dashes (—) indicate sequence identity with the consensus. Hyphens (–) indicate sequence not covered by any identified peptide. **(C)** Global graphical view of (A) and (B) alignments. The first line shows the consensus sequence with the CH1, Hinge, CH2, and CH3 domains highlighted in light green, dark blue, dark green, and light blue, respectively. Mutation sites are indicated in black above the consensus, with the position and nature of the substituted amino acids. For each allele, the detected and undetected sequences are color-coded as follows: orange = sequence detected from the LysC proteolytic mix; purple = sequence detected from the Trypsin+LysC proteolytic mix; red = sequence detected in both proteolytic mixes; dark grey = undetected sequence; light grey = sequence absent in this allele. Allele nomenclature follows the IMGT database (<https://imgt.org/>).
