## Supplementary material for "Isolation of *Toxoplasma gondii*-specific IgG and mass spectrometric characterization of individual IgGs for early diagnosis of toxoplasmosis": S4_Fig

### **S4 Fig. DIA MS/MS spectrum confirming the detection of the IGHG1*07 peptide.**

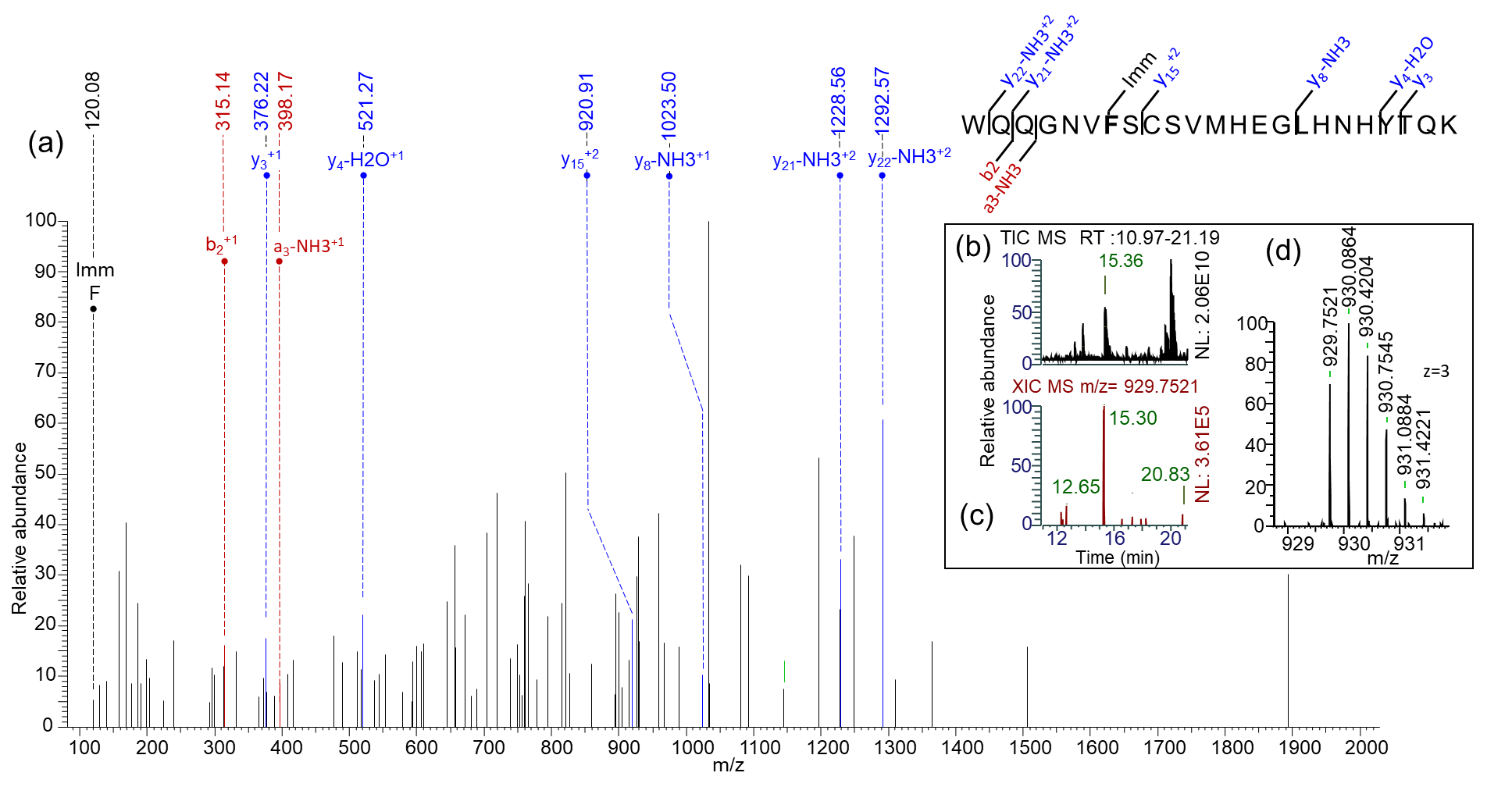

MS and DIA MS/MS evidence for the immunoglobulin heavy chain peptide WQQGNVFSCSVMHEGLHNHYTQK from IGHG01*07: **(a)** Higher-energy collision dissociation (HCD) DIA spectrum of the precursor ion m/z 929.7521 (z = 3), showing fragment ions matching the theoretical peptide sequence WQQGNVFSCSVMHEGLHNHYTQK within a 30 ppm mass tolerance and annotated b- and y-series ions; **(b)** Total ion chromatogram (TIC) (RT: 10.97–21.19 min); **(c)** EXtracted ion chromatogram (XIC) of the precursor ion m/z 929.7521 highlighting the retention time 15.30 min; **(d)** Full scan MS enlarged isotopic distribution of the precursor ion confirming the triply charged species (z = 3).
