## Supplementary material for "Isolation of *Toxoplasma gondii*-specific IgG and mass spectrometric characterization of individual IgGs for early diagnosis of toxoplasmosis": S1_Table

### **S1 Table. Amino acid variations in the CH1, CH2 and CH3-CHS constant domains, and in the Hinge regions of IGHG**

**S1-1 Table.** Amino acid variations in the CH1 constant domain of IGHG.

**S1-2 Table.** Amino acid variations in hinge regions of IGHG.

**S1-3 Table.** Amino acid variations in the CH2 constant domain of IGHG.

**S1-4 Table.** Amino acid variations in the CH3-CHS constant domain of IGHG.

**S1-1 Table. Amino acid variations in the CH1 constant domain of IGHG**

| IMGT unique numbering for C-DOMAIN [1] | 10 | 12 | 16 | 17 | 79 | 84.3 | 88 | 92 | 95 | 96 | 100 | 103 | 107 | 120 |
| --- | --- | --- | --- | --- | --- | --- | --- | --- | --- | --- | --- | --- | --- | --- |
| EU numbering [2] | 131 | 133 | 137 | 138 | 168 | 176 | 185 | 189 | 192 | 193 | 196 | 199 | 203 | 214 |
| IMGT IGHG1*01... / pmIG IGHG1_01...^a^ | **S** | **K** | **G** | **G** | H | S | V | P | **S** | L | **Q** | **I** | **N** | K |
| IMGT IGHG1*03... / pmIG IGHG1_03...^b^ |  |  |  |  |  |  |  |  |  |  |  |  |  | **R** |
| pmIG IGHG1_17_20 |  |  |  |  |  |  |  |  |  |  |  | T |  | **R** |
| pmIG IGHG1_13 | C |  |  |  |  |  |  |  |  |  |  |  |  | **R** |
| IMGT IGHG2*01... / pmIG IGHG2_02...^c^ | **C** | **R** | **E** | **S** | H | S | V | P | **N** | F | **Q** | **T** | **D** | **T** |
| IMGT IGHG2*09 |  |  | G | G |  |  |  |  |  | L |  |  |  |  |
| IMGT IGHG2*02 |  |  | G | G |  |  |  | T |  | L |  |  |  |  |
| IMGT IGHG2*04 |  |  | G | G |  |  |  |  | S | L |  |  |  |  |
| pmIG IGHG2_03 |  |  |  |  |  |  |  |  | S | L |  |  |  |  |
| pmIG IGHG2_01_06_11 |  |  |  |  |  |  |  | T |  | F |  |  |  |  |
| pmIG IGHG2_17 |  |  |  |  |  |  | M | S |  | F |  |  |  |  |
| IMGT IGHG3*01... / pmIG IGHG3_01…^d^ | **C** | **R** | **G** | **G** | H | S | V | P | **S** | L | **Q** | **T** | **N** | **R** |
| IMGT IGHG3*02 | - | - | - | - | - | - | - | - | - | - | - | - | - |  |
| IMGT IGHG3*18 / pmIG IGHG3_07_28 |  |  |  |  |  | Y |  |  |  |  |  |  |  |  |
| IMGT IGHG3*17 / pmIG IGHG3_06_40 |  |  |  |  |  |  |  |  | N | F |  |  |  |  |
| pmIG IGHG3_17_30 |  |  |  |  |  |  |  |  |  |  |  | I |  |  |
| IMGT IGHG4*01... / pmIG IGHG4_01…^e^ | **C** | **R** | **E** | **S** | H | S | V | P | **S** | L | **K** | **T** | **D** | **R** |
| pmIG IGHG4_18 |  |  |  |  | Q |  |  |  |  |  |  |  |  |  |
| pmIG IGHG4_38 |  |  |  |  |  |  |  |  |  |  | Q |  | N |  |
| pmIG IGHG4_43 |  |  |  |  |  |  |  |  |  |  | Q |  |  |  |

Reference sequences are represented by allele 01 of each IGHG subclasses following IMGT® (<https://www.imgt.org/>)

The IGHG subclasses names coming from IMGT® are write in black with * for indicating allele numbering; these come from pmIG [3] are write in blue and alleles are named with _

In bold (black, red or green): at the given position, polymorphism between subclasses; column with black and red bold: only one subclass (red bold) is different from the other three (black bold); column with black and green bold: two subclasses have the same variation.

Highlighted in yellow: at the given position AA considered as variation in the subclass where is yellow highlighted but existing (and normal) in other subclasses.

Highlighted in red: at the given position one-off modification of an AA.

IMGT IGHG2/CH1: there is no *16 and *17 (appear at CH2); *18 is non-existent.

IMGT IGHG3*02/CH1: start of nucleotide sequence/prot towards the end of CH1.

IMGT IGHG4/CH1: there is no *02, *03, *05 and *06 (appear either at the Hinge or at the CH2 level); *08 is non-existent.

^a^. equivalent AA sequence of CH1 domain between IMGT IGHG1*01*02*04*05*07*09*10*11*12*13*14*15*16 and pmIG IGHG1_01_02_05_07_09_10_11_12_14_16_18_19_21

^b^. equivalent AA sequence of CH1 domain between IMGT IGHG1*03*06*08*17 and pmIG IGHG1_03_04_06_08_15

^c^. equivalent AA sequence of CH1 domain between IMGT IGHG2*01*03*05*06*07*08*10*11*12*13*14*15*19 and pmIG IGHG2_02_04_05_07_08_09_10_12_13_14_15_16_18_19_20

^d^. equivalent AA sequence of CH1 domain between IMGT IGHG3*01*03 to *16 and *19 to *29 and pmIG IGHG3_01 to _05 and _08 to _16 and _18 to _27_29 and _31 to _39

^e^. equivalent AA sequence of CH1 domain between IMGT IGHG4*01*04*07*09*10 ; IGHG4A*01*02 and pmIG IGHG4_01 to _17 then _19 to_37 then 39_40_41_42_44

**S1-2 Table. Amino acid variations in hinge regions of IGHG**

| Hinge domain | Hinge 1 | | | | | | | | | | | | | | | Hinge 2 | | | | | | | | | | | | | | | | | Hinge 3 | | | | | | | | | | | | | | | Hinge 4 | | | | | | | | | | | | | | |
| --- | --- | --- | --- | --- | --- | --- | --- | --- | --- | --- | --- | --- | --- | --- | --- | --- | --- | --- | --- | --- | --- | --- | --- | --- | --- | --- | --- | --- | --- | --- | --- | --- | --- | --- | --- | --- | --- | --- | --- | --- | --- | --- | --- | --- | --- | --- | --- | --- | --- | --- | --- | --- | --- | --- | --- | --- | --- | --- | --- | --- | --- | --- |
| IMGT unique numbering for C-DOMAIN[1] | 1 | 2 | 3 | 4 | 5 | 6 | 7 | 8 | 9 | 10 | 11 | 12 | 13 | 14 | 15 |  |  |  |  |  |  |  |  |  |  |  |  |  |  |  |  |  |  |  |  |  |  |  |  |  |  |  |  |  |  |  |  |  |  |  |  |  |  |  |  |  |  |  |  |  |  |  |
| EU numbering[2] | 216 | 217 | 218 | 219 | 220 | 221 | 222 | 223 | 224 | 225 | 226 | 227 | 228 | 229 | 230 |  |  |  |  |  |  |  |  |  |  |  |  |  |  |  |  |  |  |  |  |  |  |  |  |  |  |  |  |  |  |  |  |  |  |  |  |  |  |  |  |  |  |  |  |  |  |  |
| IMGT IGHG1*01 to *17 / pmIG IGHG1_01 to _21 | E | P | K | S | C | D | K | T | H | T | C | P | P | C | P |  |  |  |  |  |  |  |  |  |  |  |  |  |  |  |  |  |  |  |  |  |  |  |  |  |  |  |  |  |  |  |  |  |  |  |  |  |  |  |  |  |  |  |  |  |  |  |
| IMGT unique numbering for C-DOMAIN[1] | 1 | 2 | 3 | 4 | 5 | 6 | 7 | 8 | 9 | 10 | 11 | 12 |  |  |  |  |  |  |  |  |  |  |  |  |  |  |  |  |  |  |  |  |  |  |  |  |  |  |  |  |  |  |  |  |  |  |  |  |  |  |  |  |  |  |  |  |  |  |  |  |  |  |
| EU numbering[2] | 216 | 217 | 218 | 219 | 220 | 222 | 224 | 226 | 227 | 228 | 229 | 230 |  |  |  |  |  |  |  |  |  |  |  |  |  |  |  |  |  |  |  |  |  |  |  |  |  |  |  |  |  |  |  |  |  |  |  |  |  |  |  |  |  |  |  |  |  |  |  |  |  |  |
| IMGT IGHG2*01* to *19 / pmIG IGHG2_01 to _20 | E | R | K | C | C | V | E | C | P | P | C | P |  |  |  |  |  |  |  |  |  |  |  |  |  |  |  |  |  |  |  |  |  |  |  |  |  |  |  |  |  |  |  |  |  |  |  |  |  |  |  |  |  |  |  |  |  |  |  |  |  |  |
| IMGT unique numbering for C-DOMAIN[1] | 1 | 2 | 3 | 4 | 5 | 6 | 7 | 8 | 9 | 10 | 11 | 12 | 13 | 14 | 15 | 16 | 17 | 18 | 19 | 20 | 21 | 22 | 23 | 24 | 25 | 26 | 27 | 28 | 29 | 30 | 31 | 32 | 33 | 34 | 35 | 36 | 37 | 38 | 39 | 40 | 41 | 42 | 43 | 44 | 45 | 46 | 47 | 48 | 49 | 50 | 51 | 52 | 53 | 54 | 55 | 56 | 57 | 58 | 59 | 60 | 61 | 62 |
| EU numbering[2] | 216 | 217 | 218 | - | - | 219 | 220 | 221 | 222 | 223 | 224 | 225 | 226 | 227 | 228 | - | - | - | - | - | - | - | - | - | - | - | - | - | - | - | - | - | - | - | - | - | - | - | - | - | - | - | - | - | - | - | - | - | - | - | - | - | - | - | - | - | - | - | - | - | 229 | 230 |
| IMGT IGHG3*01*02*05 to *11*13*14*15*16*20*21*22*24 to *29 / pmIG IGHG3_01 to _40 | E | L | K | T | P | L | G | D | T | T | H | T | C | P | R | C | P | E | P | K | S | C | D | T | P | P | P | C | P | R | C | P | E | P | K | S | C | D | T | P | P | P | C | P | R | C | P | E | P | K | S | C | D | T | P | P | P | C | P | R | C | P |
| IMGT IGHG3*03*17*18*19 (H1+H3+H4) |  |  |  |  |  |  |  |  |  |  |  |  |  |  |  |  |  |  |  |  |  |  |  |  |  |  |  |  |  |  |  |  |  |  |  |  |  |  |  |  |  |  |  |  |  |  |  | - | - | - | - | - | - | - | - | - | - | - | - | - | - | - |
| IMGT IGHG3*12*23 (H1+H2+H4) |  |  |  |  |  |  |  |  |  |  |  |  |  |  |  |  |  |  |  |  |  |  |  |  |  |  |  |  |  |  |  |  |  |  |  |  |  |  |  |  |  |  |  |  |  |  |  | - | - | - | - | - | - | - | - | - | - | - | - | - | - | - |
| IMGT IGHG3*04 (H1+H4) |  |  |  |  |  |  |  |  |  |  |  |  |  |  |  |  |  |  |  |  |  |  |  |  |  |  |  |  |  |  |  |  | - | - | - | - | - | - | - | - | - | - | - | - | - | - | - | - | - | - | - | - | - | - | - | - | - | - | - | - | - | - |
| IMGT unique numbering for C-DOMAIN[1] | 1 | 2 | 3 | 4 | 5 | 6 | 7 | 8 | 9 | 10 | 11 | 12 |  |  |  |  |  |  |  |  |  |  |  |  |  |  |  |  |  |  |  |  |  |  |  |  |  |  |  |  |  |  |  |  |  |  |  |  |  |  |  |  |  |  |  |  |  |  |  |  |  |  |
| EU numbering[2] | 216 | 217 | 218 | - | - | 224 | 225 | 226 | 227 | 228 | 229 | 230 |  |  |  |  |  |  |  |  |  |  |  |  |  |  |  |  |  |  |  |  |  |  |  |  |  |  |  |  |  |  |  |  |  |  |  |  |  |  |  |  |  |  |  |  |  |  |  |  |  |  |
| IMGT IGHG4*01*02*04*07*09*10 ; IGHG4A*01*02 / pmIG IGHG4_01 to _44 (except_11) | E | S | K | Y | G | P | P | C | P | S | C | P |  |  |  |  |  |  |  |  |  |  |  |  |  |  |  |  |  |  |  |  |  |  |  |  |  |  |  |  |  |  |  |  |  |  |  |  |  |  |  |  |  |  |  |  |  |  |  |  |  |  |
| pmIG IGHG4_11 |  | P |  |  |  |  |  |  |  |  |  |  |  |  |  |  |  |  |  |  |  |  |  |  |  |  |  |  |  |  |  |  |  |  |  |  |  |  |  |  |  |  |  |  |  |  |  |  |  |  |  |  |  |  |  |  |  |  |  |  |  |  |

Reference sequences are represented by allele 01 of each IGHG subclasses following IMGT® (<https://www.imgt.org/>)

The IGHG subclasses names coming from IMGT® are write in black with * for indicating allele numbering; these come from pmIG [3] are write in blue and alleles are named with _

IMGT IGHG2/Hinge : there is no *16 and *17 (appear at CH2); *18 is non-existent.

IMGT IGHG3/Hinge : be careful about IMGT numbering, there is a difference between "/IMGT repertoire/Proteins/alleles/" and "IMGTScientificChart/Numbering/"

IMGT IGHG4/Hinge : there is no *03*05 and *06 (appear at CH2 level) ; *08 is non-existent.

Highlighted in yellow: at the given position AA considered as variation in the subclass where is yellow highlighted but existing (and normal) in other subclasses.

In transparent: HINGE 1

Highlighted in orange : HINGE 2

Highlighted in blue : HINGE 3

Highlighted in purple : HINGE 4

**S1-3 Table. Amino acid variations in the CH2 constant domain of IGHG**

| IMGT unique numbering for C-DOMAIN[1] | 1.4 | 1.3 | 1.2 | 1.1 | 1 | 2 | 3 | 7 | 15.2 | 27 | 28 | 29 | 30 | 31 | 34 | 38 | 40 | 44 | 45.1 | 82 | 83 | 84 | 84.2 | 84.3 | 84.4 | 85.4 | 85.3 | 85.2 | 85.1 | 86 | 88 | 91 | 92 | 93 | 105 | 107 | 108 | 109 | 110 | 113 | 114 | 115 | 116 | 117 | 118 | 119 | 123 | 124 |
| --- | --- | --- | --- | --- | --- | --- | --- | --- | --- | --- | --- | --- | --- | --- | --- | --- | --- | --- | --- | --- | --- | --- | --- | --- | --- | --- | --- | --- | --- | --- | --- | --- | --- | --- | --- | --- | --- | --- | --- | --- | --- | --- | --- | --- | --- | --- | --- | --- |
| EU numbering[2] | 233 | 234 | 235 | 236 | 237 | 238 | 239 | 243 | 253 | 265 | 266 | 267 | 268 | 269 | 270 | 274 | 276 | 280 | 282 | 291 | 292 | 293 | 295 | 296 | 297 | 298 | 299 | 300 | 301 | 303 | 305 | 308 | 309 | 310 | 322 | 324 | 325 | 326 | 327 | 328 | 329 | 330 | 331 | 332 | 333 | 334 | 338 | 339 |
| IMGT IGHG1*01... / pmIG IGHG1_01...^a^ | E | L | L | G | G | P | S | F | I | D | V | S | **H** | E | D | **K** | **N** | D | V | P | R | E | Q | **Y** | N | S | T | **Y** | R | V | V | V | **L** | H | K | S | N | K | **A** | L | P | A | **P** | I | E | K | K | **A** |
| IMGT IGHG1*07 / pmIG IGHG1_05 |  |  |  |  |  |  |  |  |  |  |  |  |  |  |  |  |  |  |  |  |  |  |  |  |  |  |  |  |  |  |  |  |  |  |  |  |  |  |  |  |  |  |  |  |  |  |  |  |
| IMGT IGHG1*04 |  |  |  |  |  |  |  |  |  |  |  |  |  |  |  |  |  |  |  |  |  |  |  |  |  |  |  |  |  |  |  |  |  |  |  |  |  |  |  |  |  |  |  |  |  |  |  |  |
| IMGT IGHG1*03*06 / pmIG IGHG1_03_09_13_15_20 |  |  |  |  |  |  |  |  |  |  |  |  |  |  |  |  |  |  |  |  |  |  |  |  |  |  |  |  |  |  |  |  |  |  |  |  |  |  |  |  |  |  |  |  |  |  |  |  |
| IMGT IGHG1*11 |  |  |  |  |  |  |  |  |  |  |  |  |  |  |  |  |  |  |  |  |  |  |  |  |  |  |  |  |  |  |  |  | V |  |  |  |  |  |  |  |  |  |  |  |  |  |  |  |
| IMGT IGHG1*13 |  |  |  |  |  |  |  |  |  |  |  |  |  |  |  |  |  |  |  |  |  |  |  | F |  |  |  |  |  |  |  |  |  |  |  |  |  |  |  |  |  |  |  |  |  |  |  |  |
| pmIG IGHG1_11 |  |  |  |  |  |  |  |  |  |  |  |  |  |  |  |  |  |  |  |  |  |  |  |  |  |  |  |  |  |  |  |  |  |  |  |  |  |  |  |  |  |  |  |  |  |  |  |  |
| pmIG IGHG1_18 |  |  |  |  |  |  |  |  |  |  |  |  |  |  |  |  |  | E |  |  |  |  |  |  |  |  |  |  |  |  |  |  |  |  |  |  |  |  |  |  |  |  |  |  |  |  |  |  |
| IMGT IGHG2*01... / pmIG IGHG2_02...^b^ | - | P | V | A | G | P | S | F | I | D | V | S | **H** | E | D | **Q** | **N** | D | V | P | R | E | Q | **F** | N | S | T | **F** | R | V | V | V | **V** | H | K | S | N | K | **G** | L | P | A | **P** | I | E | K | K | **T** |
| IMGT IGHG2*15 / pmIG IGHG2_15 |  |  |  |  |  |  |  |  |  |  |  |  |  |  |  |  |  |  |  |  |  |  |  |  |  |  |  |  |  |  |  |  |  |  |  |  |  |  |  |  |  |  |  |  |  |  |  |  |
| IMGT IGHG2*06 / pmIG IGHG2_04_20 |  |  |  |  |  |  |  |  |  |  |  |  |  |  |  |  |  |  |  |  |  |  |  |  |  |  |  |  |  |  |  |  |  |  |  |  |  |  |  |  |  |  |  |  |  |  |  |  |
| IMGT IGHG2*11*14 / pmIG IGHG2_16 |  |  |  |  |  |  |  |  |  |  |  |  |  |  |  |  |  |  |  |  |  |  |  |  |  |  |  |  |  |  |  |  | L |  |  |  |  |  |  |  |  |  |  |  |  |  |  |  |
| IMGT IGHG2*02 / pmIG IGHG2_01_05_11 |  |  |  |  |  |  |  |  |  |  |  |  |  |  |  |  |  |  | M |  |  |  |  |  |  |  |  |  |  |  |  |  |  |  |  |  |  |  |  |  |  |  |  |  |  |  |  |  |
| pmIG IGHG2_17 |  |  |  |  |  |  |  |  |  |  |  |  |  |  |  |  |  |  |  |  |  |  |  |  |  |  |  |  |  |  |  |  |  |  |  |  |  |  |  |  |  |  |  |  |  |  |  |  |
| IMGT IGHG3*01... / pmIG IGHG3_01…^c^ | E | L | L | G | G | P | S | F | I | D | V | S | **H** | E | D | **Q** | **K** | D | V | P | R | E | Q | **Y** | N | S | T | **F** | R | V | V | V | **L** | H | K | S | N | K | **A** | L | P | A | **P** | I | E | K | K | **T** |
| IMGT IGHG3*03 |  |  |  |  |  |  |  |  |  |  |  |  |  |  |  |  |  |  |  |  |  |  |  |  |  |  |  |  |  |  |  |  |  |  |  |  |  |  |  |  |  |  |  |  |  |  |  |  |
| IMGT IGHG3*06*07 / pmIG IGHG3_02_18 |  |  |  |  |  |  |  |  |  |  |  |  |  |  |  |  |  |  |  |  |  |  |  |  |  |  |  |  |  |  |  |  |  |  |  |  |  |  |  |  |  |  |  |  |  |  |  |  |
| IMGT IGHG3*13 |  |  |  |  |  |  |  |  |  |  |  |  |  |  |  |  |  |  |  |  |  |  |  |  |  |  |  |  |  |  |  |  |  |  |  |  |  |  |  |  |  |  |  |  |  |  |  |  |
| IMGT IGHG3*08 / pmIG IGHG3_03_39 |  |  |  |  |  |  |  |  |  |  |  |  |  |  |  |  |  |  |  |  |  |  |  |  |  |  |  |  |  |  |  |  |  |  |  |  |  |  |  |  |  |  |  |  |  |  |  |  |
| IMGT IGHG3*24 |  |  |  |  |  |  |  |  |  |  |  |  |  |  |  |  |  |  |  |  |  |  |  |  |  |  |  |  |  |  |  |  |  |  |  |  |  |  |  |  |  |  |  |  |  |  |  |  |
| IMGT IGHG3*17 / pmIG IGHG3_06 |  |  |  |  |  |  |  |  |  |  |  |  |  |  |  |  |  |  |  |  |  |  |  |  |  |  |  |  |  |  |  |  |  |  |  |  |  |  |  |  |  |  |  |  |  |  |  |  |
| IMGT IGHG3*09 / pmIG IGHG3_04 |  |  |  |  |  |  |  |  |  |  |  |  |  |  |  |  |  |  |  |  |  |  |  |  |  |  |  |  |  |  |  |  | V |  |  |  |  |  |  |  |  |  |  |  |  |  |  |  |
| IMGT IGHG3*11*12*28 / pmIG IGHG3_09 |  |  |  |  |  |  |  |  |  |  |  |  |  |  |  |  |  |  |  |  |  |  |  | F |  |  |  |  |  |  |  |  |  |  |  |  |  |  |  |  |  |  |  |  |  |  |  |  |
| IMGT IGHG3*26 |  |  |  |  |  |  |  |  |  |  |  |  |  |  |  |  |  |  |  |  |  |  |  | F |  |  |  |  |  |  |  |  |  |  |  |  |  |  | G |  |  |  |  |  |  |  |  |  |
| IMGT IGHG3*18*19 / pmIG IGHG3_07_08_26_28 |  |  |  |  |  |  |  |  |  |  |  |  |  |  |  |  |  |  |  |  | W |  |  |  |  |  |  |  |  |  |  |  |  |  |  |  |  |  |  |  |  |  |  |  |  |  |  |  |
| IMGT IGHG3*23 |  |  |  |  |  |  |  |  |  |  |  |  |  |  |  |  |  |  |  |  | W |  |  |  |  |  |  |  |  |  |  |  |  |  |  |  |  |  |  |  |  |  |  |  |  |  |  |  |
| IMGT IGHG3*14*27*29 / pmIG IGHG3_05 |  |  |  |  |  |  |  |  |  |  |  |  |  |  |  |  |  |  |  | L |  |  |  |  |  |  |  |  |  |  |  |  |  |  |  |  |  |  |  |  |  |  |  |  |  |  |  |  |
| IMGT IGHG3*22 / pmIG IGHG3_19 |  |  |  |  |  |  |  |  |  |  |  |  |  |  |  |  |  |  |  | L |  |  |  |  |  |  |  |  |  |  |  |  |  |  |  |  |  |  |  |  |  |  |  |  |  |  |  |  |
| IMGT IGHG3*15 |  |  |  |  |  |  |  |  |  |  |  |  |  |  |  |  |  |  |  | L |  |  |  |  |  |  |  |  |  |  |  |  |  |  |  |  |  |  |  |  |  |  |  |  |  |  |  |  |
| IMGT IGHG3*20 |  |  |  |  |  |  |  |  |  |  |  |  |  |  |  |  |  |  |  | L |  |  |  |  |  |  |  |  |  |  |  |  |  |  |  |  |  |  |  |  |  |  |  |  |  |  |  |  |
| IMGT IGHG3*16 / pmIG IGHG3_23 |  |  |  |  |  |  |  |  |  |  |  |  |  |  |  |  |  |  |  | L |  |  |  |  |  |  |  |  |  |  |  |  |  |  |  |  |  |  |  |  |  |  |  |  |  |  |  | A |
| IMGT IGHG3*25 / pmIG IGHG3_32 |  |  |  |  |  |  |  |  |  |  |  |  |  |  |  | K |  |  |  | L |  |  |  |  |  |  |  |  |  |  |  |  |  |  |  |  |  |  |  |  |  |  |  |  |  |  |  |  |
| pmIG IGHG3_12 |  |  |  |  |  |  |  |  |  |  |  |  |  |  |  |  |  |  |  | L |  |  |  |  |  |  |  |  |  |  |  |  |  |  |  |  |  |  |  |  |  |  |  |  |  |  |  |  |
| pmIG IGHG3_13 |  |  |  |  |  |  |  |  |  |  |  |  |  |  |  |  |  |  |  |  |  |  |  |  |  |  |  |  |  |  |  |  |  |  |  |  |  |  |  |  |  |  |  |  |  |  |  |  |
| pmIG IGHG3_14 |  |  |  |  |  |  |  |  |  |  |  |  |  |  |  |  |  |  |  |  |  |  |  | F |  |  |  |  |  |  |  |  |  |  |  |  |  |  |  |  |  |  |  |  |  |  |  |  |
| pmIG IGHG3_15 |  |  |  |  |  |  |  |  |  |  |  |  |  |  |  |  |  |  |  |  |  |  |  | F |  |  |  |  |  |  |  |  |  |  |  |  |  |  |  |  |  |  |  |  |  |  |  |  |
| pmIG IGHG3_16_30_31 |  |  |  |  |  |  |  |  |  |  |  |  |  |  |  |  |  |  |  |  |  |  |  |  |  |  |  |  |  |  |  |  |  |  |  |  |  |  |  |  |  |  |  |  |  |  |  |  |
| pmIG IGHG3_20_34 |  |  |  |  |  |  |  |  |  |  |  |  |  |  |  |  |  |  |  |  |  |  |  |  |  |  |  |  |  |  |  |  |  |  |  |  |  |  |  |  |  |  |  |  |  |  |  |  |
| pmIG IGHG3_21 |  |  |  |  |  |  |  |  |  |  |  |  |  |  |  |  |  |  |  |  |  |  |  |  |  |  |  |  |  |  |  |  |  |  |  |  |  |  |  |  |  |  |  |  |  |  |  |  |
| pmIG IGHG3_22 |  |  |  |  |  |  |  |  |  |  |  |  |  |  |  |  |  |  |  |  |  |  |  | F |  |  |  |  |  |  |  |  |  |  |  |  |  |  |  |  |  |  |  |  |  |  |  |  |
| pmIG IGHG3_24 |  |  |  |  |  |  |  |  |  |  |  |  |  |  |  |  |  |  |  |  |  |  |  |  |  |  |  |  |  |  |  |  |  |  |  |  |  |  |  |  |  |  |  |  |  |  |  |  |
| pmIG IGHG3_25 |  |  |  |  |  |  |  |  |  |  |  |  |  |  |  |  |  |  |  |  |  |  |  | F |  |  |  |  |  |  |  |  |  |  |  |  |  |  |  |  |  |  |  |  |  |  |  |  |
| pmIG IGHG3_27 |  |  |  |  |  |  |  |  |  |  |  |  |  |  |  |  |  |  |  |  |  |  |  |  |  |  |  |  |  |  |  |  |  |  |  |  |  |  |  |  |  |  |  |  |  |  |  |  |
| pmIG IGHG3_29 |  |  |  |  |  |  |  |  |  |  |  |  |  |  |  |  |  |  |  |  |  |  |  |  |  |  |  |  |  |  |  |  |  |  |  |  |  |  |  |  |  |  |  |  |  |  |  |  |
| pmIG IGHG3_33_36 |  |  |  |  |  |  |  |  |  |  |  |  |  |  |  |  |  |  |  |  |  |  |  |  |  |  |  |  |  |  |  |  |  |  |  |  |  |  |  |  |  |  |  |  |  |  |  |  |
| pmIG IGHG3_35 |  |  |  |  |  |  |  |  |  |  |  |  |  |  |  |  |  |  |  |  |  |  |  |  |  |  |  |  |  |  |  |  |  |  |  |  |  |  |  |  |  |  |  |  |  |  |  |  |
| pmIG IGHG3_37 |  |  |  |  |  |  |  |  |  |  |  |  |  |  |  |  | N |  |  |  |  |  |  |  |  |  |  |  |  |  |  |  |  |  |  |  |  |  |  |  |  |  |  |  |  |  |  |  |
| pmIG IGHG3_38 |  |  |  |  |  |  |  |  |  |  |  |  |  |  |  |  |  |  |  |  |  |  |  |  |  |  |  |  |  |  |  |  |  |  |  |  |  |  |  |  |  |  |  |  |  |  |  |  |
| pmIG IGHG3_40 |  |  |  |  |  |  |  |  |  |  |  |  |  |  |  |  |  |  |  |  |  |  |  |  |  |  |  |  |  |  |  |  |  |  |  |  |  |  |  |  |  |  |  |  |  |  |  |  |
| IMGT IGHG4*01... / pmIG IGHG4_01…^d^ | E | F | L | G | G | P | S | F | I | D | V | S | **Q** | E | D | **Q** | **N** | D | V | P | R | E | Q | **F** | N | S | T | **Y** | R | V | **V** | **V** | **L** | H | K | S | N | K | **G** | L | P | S | **S** | I | E | K | K | **A** |
| IMGT IGHG4*03*09 |  |  |  |  |  |  |  |  |  |  |  |  |  |  |  |  |  |  |  |  |  |  |  |  |  |  |  |  |  |  |  |  |  |  |  |  |  |  |  |  |  |  |  |  |  |  |  |  |
| IMGT IGHG4*02; IGHG4A*01*02/ pmIG IGHG4_03_05_06_13_20_29_43 |  |  |  |  |  |  |  |  |  |  |  |  |  |  |  |  |  |  |  |  |  |  |  |  |  |  |  |  |  |  |  |  | V |  |  |  |  |  |  |  |  |  |  |  |  |  |  |  |
| IMGT IGHG4*06 |  |  |  |  |  |  |  |  |  |  |  |  |  |  |  |  |  |  |  |  |  |  |  |  |  |  |  |  |  |  |  | I |  |  |  |  | H |  |  |  |  |  |  |  |  |  |  |  |
| IMGT IGHG4*05 / pmIG IGHG4_11 |  |  |  |  |  |  |  |  |  |  |  |  |  |  |  |  |  |  |  |  |  |  |  |  |  |  |  |  |  |  |  | I |  |  |  |  | H |  |  |  |  |  |  |  |  |  |  |  |
| pmIG IGHG4_08_25 |  |  |  |  |  |  |  |  |  |  |  |  |  |  |  |  |  |  |  |  |  |  |  |  |  |  |  |  |  |  |  |  | V |  |  |  |  |  |  |  |  |  |  |  |  |  |  |  |
| pmIG IGHG4_10_33 |  |  |  |  |  |  |  |  |  |  |  |  |  |  |  |  |  |  |  |  |  |  |  |  |  |  |  |  |  |  |  |  |  |  |  |  |  |  |  |  |  |  |  |  |  |  |  |  |
| pmIG IGHG4_15_17 |  |  |  |  |  |  |  |  |  |  |  |  |  |  |  |  |  |  |  |  |  |  |  |  |  |  |  |  |  |  |  |  |  |  |  |  |  |  |  |  |  |  |  |  |  |  |  |  |
| pmIG IGHG4_19 |  |  |  |  |  |  |  |  |  |  |  |  |  |  |  |  |  |  |  |  |  |  |  |  |  |  |  |  |  |  |  |  |  |  |  |  |  |  |  |  |  |  |  |  |  |  |  |  |
| pmIG IGHG4_21 |  |  |  |  |  |  |  |  |  |  |  |  |  |  |  |  |  |  |  |  |  |  |  |  |  |  |  | C |  |  |  |  | V |  |  |  |  |  |  |  |  |  |  |  |  |  |  |  |
| pmIG IGHG4_22 |  |  |  |  |  |  |  |  |  |  |  |  |  |  |  |  |  |  |  |  |  |  |  |  |  |  |  |  |  |  |  |  |  |  |  |  |  |  |  |  |  |  |  |  |  |  |  |  |
| pmIG IGHG4_24 |  |  |  |  |  |  |  |  |  |  |  |  |  |  |  |  |  |  |  |  |  |  |  |  |  |  |  | C |  |  |  |  | V |  |  |  |  |  |  |  |  |  |  |  |  |  |  |  |
| pmIG IGHG4_30 |  |  |  |  |  |  |  |  |  |  |  |  |  |  |  |  |  |  |  |  |  |  |  |  |  |  |  |  |  |  | I |  |  |  |  |  |  |  |  |  |  |  |  |  |  |  |  |  |
| pmIG IGHG4_35 |  |  |  |  |  |  |  |  |  |  |  |  | H |  |  |  |  |  |  |  |  |  |  |  |  |  |  |  |  |  |  |  | V |  |  |  |  |  |  |  |  |  |  |  |  |  |  |  |
| pmIG IGHG4_39 |  |  |  |  |  |  |  |  |  |  |  |  | H |  |  |  |  |  |  |  |  |  |  |  |  |  |  |  |  |  |  |  |  |  |  |  |  |  |  |  |  |  |  |  |  |  |  |  |
| pmIG IGHG4_40 |  |  |  |  |  |  |  |  |  |  |  |  |  |  |  |  |  |  |  |  |  |  |  |  |  |  |  |  |  |  |  | I |  |  |  |  | S |  | **-** |  |  |  |  |  |  |  |  |  |
| pmIG IGHG4_42 |  |  |  |  |  |  |  |  |  |  |  |  |  |  |  |  |  |  |  |  |  |  |  |  |  |  |  |  |  |  |  |  |  |  |  |  |  |  |  |  |  |  |  |  |  |  |  | T |
| pmIG IGHG4_44 |  |  |  |  |  |  |  |  |  |  |  |  |  |  |  |  |  |  |  |  |  |  |  |  |  |  |  |  |  |  |  |  |  |  |  |  |  |  |  |  |  |  |  |  |  |  |  |  |

Reference sequences are represented by allele 01 of each IGHG subclasses following IMGT® (<https://www.imgt.org/>)

The IGHG subclasses names coming from IMGT® are write in black with * for indicating allele numbering; these come from pmIG [3] are write in blue and alleles are named with _

In bold (black, red or green): at the given position, polymorphism between subclasses; column with black and red bold: only one subclass (red bold) is different from the other three (black bold); column with black and green bold: two subclasses have the same variation

Highlighted in yellow: at the given position AA considered as variation in the subclass where is yellow highlighted but existing (and normal) in other subclasses

Highlighted in red: at the given position one-off modification of an AA

Numbering highlighted in orange: glycosylation site position

IMGT IGHG2/CH2-CH3-CHS : *18 is non-existent

IMGT IGHG4/CH2-CH3-CHS : *08 is non-existent

^a^. equivalent AA sequence of CH2 domain between IMGT IGHG1*01*02*05*08*09*10*12*14*15*16*17 and pmIGHG1_01_02_04_06_07_08_10_12_14_16_17_19_21

^b^. equivalent AA sequence of CH2 domain between IMGT IGHG2*01*03*04*05*07*08*09*10*12*13*16*17*19 and pmIG IGHG2_02_03_06_07_08_09_10_12_13_14_18_19

^c^. equivalent AA sequence of CH2 domain between IMGT IGHG3*01*02*04*05*10*21 and pmIG IGHG3_01_10_11_17

^d^. equivalent AA sequence of CH2 domain between IMGT IGHG4*01*04*07*10 and pmIG IGHG4_01_02_04_07_09_12_14_16_18_23_26_27_28_31_32_34_36_37_38_41

**S1-4 Table. Amino acid variations in the CH3-CHS constant domain of IGHG**

| IMGT unique numbering for C-DOMAIN[1] | 8 | 11 | 12 | 14 | 36 | 38 | 39 | 44 | 45.2 | 79 | 80 | 81 | 83 | 84 | 88 | 98 | 101 | 110 | 115 | 116 | 125 |
| --- | --- | --- | --- | --- | --- | --- | --- | --- | --- | --- | --- | --- | --- | --- | --- | --- | --- | --- | --- | --- | --- |
| EU numbering[2] | 352 | 355 | 356 | 358 | 376 | 378 | 379 | 384 | 387 | 392 | 393 | 394 | 396 | 397 | 409 | 419 | 422 | 431 | 435 | 436 | 445 |
| IMGT IGHG1*01... / pmIG IGHG1_01...^a^ | P | **R** | **D** | **L** | D | A | V | **N** | P | **K** | T | T | P | **V** | **K** | **Q** | **V** | A | **H** | **Y** | **P** |
| IMGT IGHG1*07 / pmIG IGHG1_05 |  |  |  |  |  |  |  |  |  |  |  |  |  |  |  |  |  | G |  |  |  |
| IMGT IGHG1*04 |  |  |  |  |  |  |  |  |  |  |  |  |  |  |  |  | I |  |  |  |  |
| IMGT IGHG1*03*06 / pmIG IGHG1_03...^b^ |  |  | E | M |  |  |  |  |  |  |  |  |  |  |  |  |  |  |  |  |  |
| IMGT IGHG1*11 |  |  |  |  |  |  |  |  |  |  |  |  |  |  |  |  |  |  |  |  |  |
| IMGT IGHG1*13 |  |  |  |  |  |  |  |  |  |  |  |  |  |  |  |  |  |  |  |  |  |
| pmIG IGHG1_11 |  |  | E | M |  |  |  |  |  |  |  |  |  |  |  |  |  | G |  |  |  |
| pmIG IGHG1_18 |  |  |  |  |  |  |  |  |  |  |  |  |  |  |  |  |  |  |  |  |  |
| IMGT IGHG2*01... / pmIG IGHG2_02...^c^ | P | **R** | **E** | **M** | D | A | V | **N** | P | **K** | T | T | P | **M** | **K** | **Q** | **V** |  | **H** | **Y** | **P** |
| IMGT IGHG2*15 / pmIG IGHG2_15 |  |  |  |  |  |  |  |  |  | N |  |  |  |  |  |  |  |  |  |  |  |
| IMGT IGHG2*06 / pmIG IGHG2_04_20 |  |  |  |  |  | **S** |  |  |  |  |  |  |  |  |  |  |  |  |  |  |  |
| IMGT IGHG2*11*14 / pmIG IGHG2_16 |  |  |  |  |  |  |  |  |  |  |  |  |  |  |  |  |  |  |  |  |  |
| IMGT IGHG2*02 / pmIG IGHG2_01_05_11 |  |  |  |  |  |  |  |  |  |  |  |  |  |  |  |  |  |  |  |  |  |
| pmIG IGHG2_17 | T |  |  |  |  |  |  |  |  |  |  |  |  |  |  |  |  |  |  |  |  |
| IMGT IGHG3*01... / pmIG IGHG3_01…^d^ | P | **R** | **E** | **M** | D | A | V | **S** | P | **N** | T | T | P | **M** | **K** | **Q** | **I** | A | **R** | **F** | **P** |
| IMGT IGHG3*03 |  |  |  |  |  |  |  |  |  |  |  |  |  | V | R | E | V |  |  |  |  |
| IMGT IGHG3*06*07 / pmIG IGHG3_02_18 |  |  |  |  |  |  |  |  |  | K |  |  |  |  |  |  |  |  |  |  |  |
| IMGT IGHG3*13 |  |  |  |  |  |  |  |  |  | K |  |  |  |  |  | E |  |  |  |  |  |
| IMGT IGHG3*08 / pmIG IGHG3_03_39 |  |  |  |  |  |  |  | N |  |  |  |  |  |  |  |  |  |  |  |  |  |
| IMGT IGHG3*24 |  |  |  |  |  |  |  | N |  | K |  |  |  |  |  |  |  |  |  | Y |  |
| IMGT IGHG3*17 / pmIG IGHG3_06 |  |  |  |  |  |  | **M** |  |  | K |  |  |  | V |  |  |  |  | H | Y |  |
| IMGT IGHG3*09 / pmIG IGHG3_04 |  |  |  |  |  |  |  |  |  |  |  |  |  |  |  |  |  |  |  |  |  |
| IMGT IGHG3*11*12*28 / pmIG IGHG3_09 |  |  |  |  |  |  |  |  |  |  |  |  |  |  |  |  |  |  |  |  |  |
| IMGT IGHG3*26 |  |  |  |  |  |  |  |  |  |  |  |  |  |  |  |  |  |  |  |  |  |
| IMGT IGHG3*18*19 / pmIG IGHG3_07_08_26_28 |  |  |  |  |  |  | **M** |  |  | K |  |  |  | V |  |  |  |  | H | Y |  |
| IMGT IGHG3*23 |  |  |  |  |  |  |  |  |  | K |  |  |  | V |  |  |  |  | H | Y |  |
| IMGT IGHG3*14*27*29 / pmIG IGHG3_05 |  |  |  |  |  |  |  | N |  |  |  |  |  |  |  |  |  |  |  | Y |  |
| IMGT IGHG3*22 / pmIG IGHG3_19 |  |  |  |  |  |  |  | N |  |  |  |  |  |  |  |  |  |  | H | Y |  |
| IMGT IGHG3*15 |  |  |  |  |  |  |  | N |  | K |  |  |  |  |  |  |  |  |  | Y |  |
| IMGT IGHG3*20 |  |  |  |  |  |  |  | N | R |  |  |  |  |  |  |  |  |  |  | Y |  |
| IMGT IGHG3*16 / pmIG IGHG3_23 |  |  |  |  |  |  |  | N |  |  |  |  |  |  |  |  |  |  |  | Y |  |
| IMGT IGHG3*25 / pmIG IGHG3_32 |  |  |  |  |  |  |  | N |  |  |  |  |  |  |  |  |  |  |  | Y |  |
| pmIG IGHG3_12 |  |  |  |  |  |  |  |  |  |  |  |  |  |  |  |  |  |  |  | Y |  |
| pmIG IGHG3_13 |  |  |  |  |  |  |  |  |  |  |  |  |  | V | R |  | V |  |  |  |  |
| pmIG IGHG3_14 |  |  |  |  |  |  |  |  |  |  |  |  |  |  |  |  |  |  |  | Y |  |
| pmIG IGHG3_15 |  |  |  |  |  |  |  | N |  |  |  |  |  |  |  |  |  |  |  |  |  |
| pmIG IGHG3_16_30_31 |  |  |  |  |  |  |  |  |  |  |  |  |  |  |  |  |  |  |  | Y |  |
| pmIG IGHG3_20_34 |  |  |  |  |  |  |  |  |  | K |  |  |  |  |  |  |  |  |  | Y |  |
| pmIG IGHG3_21 |  |  |  |  |  |  |  |  |  |  |  |  |  | V |  |  |  |  |  |  |  |
| pmIG IGHG3_22 |  |  |  |  |  |  |  |  |  |  |  |  |  |  |  |  |  |  | H |  |  |
| pmIG IGHG3_24 |  |  |  |  |  |  |  |  |  | K |  |  |  |  |  |  |  |  | H | Y |  |
| pmIG IGHG3_25 |  |  |  |  |  |  |  |  |  |  |  |  |  |  |  |  |  |  | H | Y |  |
| pmIG IGHG3_27 |  |  |  |  |  |  |  |  |  |  |  |  |  | V |  |  | V |  |  |  |  |
| pmIG IGHG3_29 |  |  |  |  |  |  |  |  |  |  |  |  |  |  |  |  |  |  | H |  |  |
| pmIG IGHG3_33_36 |  |  |  |  |  |  |  |  |  |  |  |  |  |  | R |  | V |  |  |  |  |
| pmIG IGHG3_35 |  |  |  |  |  |  |  |  |  |  |  |  |  | V | R |  |  |  |  |  |  |
| pmIG IGHG3_37 |  |  |  |  |  |  |  |  |  |  |  |  |  |  |  |  |  |  |  |  |  |
| pmIG IGHG3_38 |  |  |  |  |  |  |  |  |  |  |  |  |  |  |  |  |  |  | H | Y |  |
| pmIG IGHG3_40 |  |  |  |  |  |  | M |  |  | K |  |  |  |  |  |  |  |  | H | Y |  |
| IMGT IGHG4*01... / pmIG IGHG4_01…^e^ | **P** | **Q** | **E** | **M** | D | A | V | **N** | P | **K** | T | T | P | **V** | **R** | **E** | **V** | A | **H** | **Y** | **L** |
| IMGT IGHG4*03*09 |  |  |  |  |  |  |  |  |  |  |  |  |  |  | K |  |  |  |  |  |  |
| IMGT IGHG4*02 ; IGHG4A*01*02 / pmIG IGHG4_03_05_06_13_20_29_43 |  |  |  |  |  |  |  |  |  |  |  |  |  |  |  |  |  |  |  |  |  |
| IMGT IGHG4*06 |  |  |  |  |  |  |  |  |  |  |  |  |  |  |  |  |  |  |  |  |  |
| IMGT IGHG4*05 / pmIG IGHG4_11 |  | R |  |  |  |  |  |  |  |  |  |  |  |  |  |  |  |  |  |  |  |
| pmIG IGHG4_08_25 |  |  |  |  |  |  |  |  |  |  |  |  |  |  |  |  |  |  |  |  | P |
| pmIG IGHG4_10_33 |  |  |  |  |  |  |  |  |  |  |  |  |  |  |  | Q | I |  |  |  | P |
| pmIG IGHG4_15_17 |  |  |  |  |  |  |  |  |  |  |  |  |  |  |  | Q | I |  |  |  |  |
| pmIG IGHG4_19 |  | P |  |  |  |  |  |  |  |  |  |  |  |  |  |  |  |  |  |  |  |
| pmIG IGHG4_21 |  |  |  |  |  |  |  |  |  |  |  |  |  |  |  |  |  |  |  |  | P |
| pmIG IGHG4_22 |  |  |  | I |  |  |  |  |  |  |  |  |  |  |  |  |  |  |  |  |  |
| pmIG IGHG4_24 |  |  |  |  |  |  |  |  |  |  |  |  |  |  |  |  |  |  |  |  |  |
| pmIG IGHG4_30 |  |  |  |  |  |  |  |  |  |  |  |  |  |  |  |  |  |  |  |  |  |
| pmIG IGHG4_35 |  |  |  |  |  |  |  |  |  |  |  |  |  |  |  |  |  |  |  |  |  |
| pmIG IGHG4_39 |  |  |  |  |  |  |  |  |  |  |  |  |  |  |  |  |  |  |  |  |  |
| pmIG IGHG4_40 |  |  |  |  |  |  |  |  |  |  |  |  |  |  |  |  |  |  |  |  |  |
| pmIG IGHG4_42 |  |  |  |  |  |  |  |  |  |  |  |  |  |  |  |  |  |  |  |  |  |
| pmIG IGHG4_44 |  |  |  |  |  |  |  |  |  |  |  |  |  |  |  |  |  |  |  | F |  |

Reference sequences are represented by allele 01 of each IGHG subclasses following IMGT® (<https://www.imgt.org/>)

The IGHG subclasses names coming from IMGT® are write in black with * for indicating allele numbering; these come from pmIG [3] are write in blue and alleles are named with _

In bold (black, red or green): at the given position, polymorphism between subclasses; column with black and red bold: only one subclass (red bold) is different from the other three (black bold); column with black and green bold: two subclasses have the same variation.

Highlighted in yellow: at the given position AA considered as variation in the subclass where is yellow highlighted but existing (and normal) in other subclasses.

Highlighted in red: at the given position one-off modification of an AA.

Numbering highlighted in orange: glycosylation site position.

IMGT IGHG2/CH2-CH3-CHS : *18 is non-existent.

IMGT IGHG4/CH2-CH3-CHS : *08 is non-existent.

^a^. equivalent AA sequence of CH3-CHS domain between IMGT IGHG1*01*02*05*08*09*10*12*14*15*16*17 and pmIGHG1_01_02_04_06_07_08_10_12_14_16_17_19_21

^b^. equivalent AA sequence of CH3-CHS domain between IMGT IGHG1*03*06 and pmIG IGHG1_03_09_13_15_20

^c^. equivalent AA sequence of CH3-CHS domain between IMGT IGHG2*01*03*04*05*07*08*09*10*12*13*16*17*19 and pmIG IGHG2_02_03_06_07_08_09_10_12_13_14_18_19

^d^. equivalent AA sequence of CH3-CHS domain between IMGT IGHG3*01*02*04*05*10*21 and pmIG IGHG3_01_10_11_17

^e^. equivalent AA sequence of CH3-CHS domain between IMGT IGHG4*01*04*07*10 and pmIG IGHG4_01_02_04_07_09_12_14_16_18_23_26_27_28_31_32_34_36_37_38_41
