## Supplementary material for "Isolation of *Toxoplasma gondii*-specific IgG and mass spectrometric characterization of individual IgGs for early diagnosis of toxoplasmosis": S2_Table

### **S2 Table. Equimolar distribution of recombinant *T. gondii* antigens used for the immunopurification columns.**

|  | ***Tg*SAG1** | ***Tg*GRA7** | ***Tg*AMA1** |
| --- | --- | --- | --- |
| Molarity weight (g/M) | 31 030 | 18 674 | 47 957 |
| Ci (mg/ml) | 1 | 2.5 | 1.5 |
| Cn (nM) | 48.34 | 48.34 | 48.34 |
| Cm (µg/ml) | 1.5 | 0.9 | 2.32 |
| Protein quantity (µg) | 15 | 9 | 23.2 |
| Beads volume (µl) | 60 | 36 | 100 |

Ci = initial concentration of the recombinant antigens; Cn = molar concentration; Cm = mass concentration. The final volume is 10 ml.
