## Supplementary material for "Isolation of *Toxoplasma gondii*-specific IgG and mass spectrometric characterization of individual IgGs for early diagnosis of toxoplasmosis": S3_Table

### **S3 Table. Molecular determination of *IGHG* alleles in the three donors: HVDtot, HVD+, and HVD-.**

|  | **HVDtot** | **HVD+** | **HVD-** |
| --- | --- | --- | --- |
| ***T. gondii* serology** | **--** | **Positive** | **Negative** |
| ***IGHG1* alleles ^a^** | *IGHG1**03 *IGHG1**07 | *IGHG1**03 *IGHG1**07 | *IGHG1**02/*08/*09/*14 (homozygous) |
| ***IGHG2* alleles ^b^** | ND | ND | ND |
| ***IGHG3* alleles ^a^** | *IGHG3**11/*12 *IGHG3**14 | *IGHG3**11/*12 *IGHG3**22 | *IGHG3**14 *IGHG3**16 |
| ***IGHG4* alleles ^b^** | ND | ND | ND |

Total IgG fractions (derived from HVDtot) and *T. gondii*-specific IgG fractions (derived from HVD+ and HVD- individuals, who differ in their serological status with respect to *T. gondii*) were used to characterize protein alleles by mass spectrometry.

- ^a^ *IGHG1* and *IGHG3* alleles were determined by molecular sequencing for all two donors. For donors with two distinct allele groups listed, each line corresponds to one allele of the heterozygous genotype.
- ^b^ ND: not determined. *IGHG2* and *IGHG4* allele assignments could not be completed due to sequence reading ambiguities (background noise). (M. Dambrun, personal communication).
- Donor codes are anonymized. Allele nomenclature follows the IMGT database (<https://imgt.org/>).
