## Supplementary material for "Isolation of *Toxoplasma gondii*-specific IgG and mass spectrometric characterization of individual IgGs for early diagnosis of toxoplasmosis": S4_Table

### **S4 Table. Mass spectrometric identification of IGHG peptides by DDA and DIA, peptide sequence, compatible allele(s), DDA Xcorr score, DIA-NN score, charge state, detection in HVD+ and HVD- samples, and validation status.**

| **Peptide sequence** | **Modification** | **Subclass** | **Compatible allele(s)** | **Domain** | **Charge** **state (z)** | **Theo. MH+** **(Da)** | **DDA Xcorr** **(Sequest HT)** | **DIA** **m/z** | **Allele** **assignment** |
| --- | --- | --- | --- | --- | --- | --- | --- | --- | --- |
| ***IGHG1 — allele-specific signature peptide (G residue, position 314)*** | | | | | | | | | |
| WQQGNVFSCSVMHE **G** LHNHYTQK | Carbamidomethyl [C9] | IGHG1 | IGHG1*07 only ^a^ | CH3 (pos. 314) | 3 | 2788.24 | 5.84 | 929.7521 | IGHG1*07 ✓ ^b^ |
| WQQGNVFSCSVMHE **G** LHNHYTQK | Carbamidomethyl [C9] | IGHG1 | IGHG1*07 only ^a^ | CH3 (pos. 314) | 5 | 2787.25 | 7.62 | — | IGHG1*07 ✓ ^b^ |
| ***IGHG1 — non-specific peptide (A residue, position 314; shared across multiple IGHG1 alleles and all IGHG2 alleles)*** | | | | | | | | | |
| WQQGNVFSCSVMHE **A** LHNHYTQK | Carbamidomethyl [C9]; Deamidated [N5] | IGHG1 / IGHG2 | IGHG1*03, *01, *02, *05, *06, *08–*17; all IGHG2 ^c^ | CH3 (pos. 314) | 3 | 2802.25 | 7.72 | — | Not assignable ^d^ |
| SRWQQGNVFSCSVMHE **A** LHNHYTQK | Carbamidomethyl [C11] | IGHG1 / IGHG2 | IGHG1*03, *01, *02, *05, *06, *08–*17; all IGHG2 ^c^ | CH3 (pos. 314) | 3 | 3044.40 | 5.85 | — | Not assignable ^d^ |
| ***IGHG3 — low-confidence candidate peptide (below DIA validation threshold)*** | | | | | | | | | |
| EEQFNSTFR | Deamidated [N5] | IGHG3 / IGHG2 | IGHG3*11; also IGHG2*17, IGHG2*02 ^e^ | CH2 (pos. 221) | 2 | 1158.51 | 3.09 | — | Not validated ^f^ |

- ^a^ The G residue at position 314 is exclusive to IGHG1*07 across the full IGHG1 allelic repertoire (IMGT database, 2024)[1]. No other IGHG1 allele carries a glycine at this position.
- ^b^ The IGHG1*07 signature peptide (G) was detected and validated in both DDA (Xcorr 5.84 at z=3 and 7.62 at z=5) and DIA (m/z 929.7521, z=3, fragment ions matched within 30 ppm tolerance). It was detected exclusively in the HVD+ seropositive sample and absent from the HVD- seronegative sample, consistent with the molecular sequencing data (see S3 Table).
- ^c^ The A residue at position 314 is present in the majority of IGHG1 alleles and in all currently referenced IGHG2 alleles. The peptide WQQGNVFSCSVMHEALHNHYTQK is therefore not allele-specific in the context of a polyclonal mixture containing both IGHG1 and IGHG2. DDA validation: Xcorr 7.72 (z=3). DIA: detected but not formally validated (insufficient fragment score profile).
- ^d^ Not assignable: the peptide is shared across multiple IGHG1 alleles and all IGHG2 alleles; individual-level or allele-level discrimination is not possible in the (HVD+)+(HVD-) mixed sample context.
- ^e^ The F residue at position 221 distinguishes IGHG3*11 from most other IGHG3 alleles, but is also present in IGHG2*17 and IGHG2*02, undermining subclass-level specificity. IGHG3*11 is the expected allele in the HVD+ donor (see Supp. Table 3).
- ^f^ Not validated: the DDA Xcorr of 3.09 (z=2) falls below the acceptance threshold for confident peptide identification. The DIA fragment score profile was insufficient for validation. This peptide was not retained for allele assignment.
- *DDA: Data-Dependent Acquisition; DIA: Data-Independent Acquisition; Xcorr: cross-correlation score (Sequest HT, Proteome Discoverer 3.2); Theo. MH+: theoretical monoisotopic mass of the singly protonated peptide; pos.: amino acid position in the IGHG protein sequence (IMGT numbering); —: not applicable or not detected.*
