## Supplementary material for "Isolation of *Toxoplasma gondii*-specific IgG and mass spectrometric characterization of individual IgGs for early diagnosis of toxoplasmosis": S1_File

S1 File. Detailed protocols for the production of recombinant *T. gondii* antigens and the characterization of IGHG alleles by mass spectrometry.

### 1 Production of *Tg*AMA1, *Tg*SAG1 and *Tg*GRA7 recombinant antigens

The techniques used for the production of *T. gondii* recombinant antigens are described elsewhere [1]. Briefly, competent SHuffle bacteria (T7 *E. coli*) for the expression of *Tg*AMA1 (67-484 aa) and *E. coli* BL21C+ for that of *Tg*SAG1 (46-336 aa) and *Tg*GRA7 (27-183 aa) were used to produce the recombinant proteins. Transformed bacteria were then cultured in appropriate conditions. After induction, an SDS-PAGE gel (Bio-Rad, France) was performed to confirm recombinant protein production at the expected molecular weights for each antigen, i.e., 50 kDa for *Tg*AMA1, 30 kDa for *Tg*SAG1 and 25 kDa for *Tg*GRA7. The bacterial cells were lysed. A solubilization step was required for *Tg*AMA1 and *Tg*SAG1. The solubilized proteins were purified on a Ni-NTA agarose resin (Fisher Scientific, France). The concentration of each eluted protein was measured at 280 nm using a Nanodrop (Thermo Fisher Scientific, France), and aliquots were stored in 10% glycerol (*Tg*AMA1, *Tg*SAG1) and 15% glycerol (*Tg*GRA7) at -80°C for long-term preservation.

### 2 Generation of the internal IGHG sequence library

An internal reference database was built for peptide identification using all available FASTA sequences corresponding to the CH2–CH3 regions of IGHG1, IGHG2, IGHG3 and IGHG4, as retrieved from the IMGT database [2]. Because proteolytic digestion can generate peptides spanning regions not included in isolated CH2–CH3 fragments — such as junctions involving CH1, the hinge (H1 to H4), or the membrane exons M1 and M2 — these peptides would not be identified if only truncated fragments were used as reference. To circumvent this limitation, full coding sequences from CH1 to M1+M2 were collected, translated into protein sequences, and incorporated into the custom reference database alongside the CH2–CH3 sequences, ensuring complete coverage of all proteolytic peptides generated across the entire IGHG constant region. One unreferenced IGHG1 allele identified in the prior proof-of-concept study was additionally included.

### 3 Mass spectrometry characterization of IGHG alleles from the Fc/2 amino-acid sequences

#### 3.1 **Middle-down analysis**

Purified IGHG alleles were detected by middle-down analysis on a Q Exactive HF Orbitrap mass spectrometer (ThermoFisher Scientific) coupled with an Ultimate 3000 nanoRSLC system. The 4 μL sample was separated on a PepMap300 C4 column (75 μm × 150 mm, 300 Å, 5 μm, ThermoFisher Scientific) at 180 nL/min using the following gradient: 0–25 min, 0–50% B; 25–30 min, 50–90% B; followed by wash and re-equilibration [mobile phases: A, 0.1% formic acid in water/acetonitrile (98:2, v/v); B, 0.1% formic acid in acetonitrile/isopropyl alcohol (80:20, v/v)]. MS data were acquired in DDA Top10 mode: 1 full MS scan at 240,000 resolution (at m/z 400) is followed by 10 HCD MS/MS scans of the top 10 precursor ions at 60,000 resolution (NCE = 20; isolation window: 30 m/z; max. inj time: 200 ms; AGC target: 1E6). Raw spectra were deconvoluted using Freestyle 1.8 SP1 software (ThermoFisher Scientific). Database searches were performed using Byonic (Protein Metrics) against our IMGT-derived IGHG sequence library, using: 10 ppm precursor mass tolerance and 20 ppm fragment mass tolerance, and considering methionine oxidation (+15.995 Da) and C-terminal lysine loss (−128.095 Da) as variable modifications. Results were filtered with the following criteria: Byonic score > 250, p-value < 10⁻⁵, delta score > 10, FDR < 1%. All retained spectral assignments were subjected to manual validation on deconvoluted MS/MS spectra [3]. Sequence coverage per allele was estimated as the ratio of unique fragment ions detected to the total number of theoretical fragments.

#### 3.2 **Optimization of Fc/2 fragment chromatographic separation**

The PepMap300 C4 nanoLC column was evaluated as a first approach for the middle-down analysis of polyclonal Fc/2 fragment mixtures. Because IGHG Fc/2 allelic variants are near-isobaric species differing by only one to a few amino acids (~14–130 Da mass difference at the ~24 kDa intact fragment level), co-elution of allelic variants on C4 conditions was assessed as a potential limitation for allele-specific precursor ion selection. To address this, an alternative large-pore reversed-phase stationary Poroshell 300SB-C3 column (5 μm, 1.0 × 75 mm, 300 Å; Agilent Technologies) was evaluated on a UHPLC Vanquish system (ThermoFisher Scientific). Analyses were performed on a UHPLC Vanquish system (ThermoFisher Scientific) at 80 μL/min and 60°C, using a gradient of solvent A (water/0.1% formic acid) and solvent B (acetonitrile/isopropyl alcohol 80:20 v/v, 0.1% formic acid) from 25% to 35% B over 10 min. Samples were loaded at 1 μg/μL after desalting on ZipTip C4. The LC was coupled to an Orbitrap Tribrid Eclipse mass spectrometer (ThermoFisher Scientific) operating at 120,000 resolution (m/z 700–2000; normalized AGC target 200%; source energy 15 V; ion transfer tube temperature 250°C; vaporizer temperature 75°C; spray voltage 3.5 kV).

### 4 Data-Dependent Acquisition (DDA) and Data-Independent Acquisition (DIA) mass spectrometry

#### 4.1 **Sample preparation for bottom-up proteomics**

We used the *T. gondii*-specific IgG fraction recovered following the Fc/2 capture step for bottom-up proteomics analysis. After in silico multi-endoprotease digestion simulation of IGHG1, IGHG2, IGHG3, and IGHG4 sequences, Trypsin, LysC, and Trypsin+LysC combinations were confirmed as the most suited ones to produce specific allele-specific proteotypic peptides. Samples were buffered with ammonium bicarbonate (pH 7.6), reduced with 5 mM DTT for 30 min at 56°C, and alkylated with 25 mM iodoacetamide at room temperature in the dark with agitation (900 rpm), before overnight proteolysis at 37°C with 2 μL of selected endoprotease(s) (0.5 μg/μL). Digestion was stopped with 1 μL of 10% TFA. Peptides were desalted using ZipTip C18 (Millipore) and eluted with 0.1% TFA in ACN/water (80:20, v/v).

#### 4.2 **NanoLC DDA and DIA CID MS/MS acquisition**

Peptide samples (2 μL) were injected onto a Neo Vanquish nanoLC system coupled to an Orbitrap Astral mass spectrometer (ThermoFisher Scientific). Peptides were separated on a 25 cm × 75 μm ID, 1.7 μm C18 Aurora Ultimate column (IonOpticks, Australia) using a 40-min gradient of solvent A (0.1% formic acid in water) and solvent B (0.1% formic acid in ACN/water (80:20, v/v)). The mass spectrometer was operated in either DDA or DIA mode. Full MS scans were acquired in the Orbitrap at 240,000 resolution, and MS/MS scans were acquired in the Astral analyzer. DDA was performed in Top100 mode with a dynamic exclusion of 15s. Sequential DIA windows were defined over the mass range 450–1500 m/z with 2 m/z isolation windows. Instrument parameters: spray voltage 2.1 kV, RF lens 30%, normalized AGC target 500%, maximum injection time 3 ms. Peptide identification was performed using Sequest™ embedded in Proteome Discoverer 3.2 and DIA-NN 1.9.1 with standard settings, for DDA and DIA analysis, respectively. Carbamidomethylation (C) was set as a fixed modification; methionine oxidation (M) and asparagine/glutamine deamidation (N, Q) were set as variable modifications, with a tolerance of 2 missed cleavages. In DIA-NN, the “duplicate-proteins” and “relaxed-prot-inf” options were enabled to account for the high sequence redundancy of the IGHG allele database. DIA-NN output was filtered to consider only IGHG identifications with at least 50% *Fragment.Correlations* values above 0. For each candidate allele-specific peptide, the raw acquisition file was re-examined at the MS1 level to verify the presence of the precursor ion at the expected charge state with a well-defined isotopic envelope. For each validated candidate peptide, the MS2 DIA spectrum was retrieved and examined for fragment ions matching the theoretical fragmentation pattern within a 30 ppm tolerance, with particular attention to fragments covering the polymorphic position.
